# Task-State Decoders as Individual-Differences Measures: Evidence from a Working Memory Neural Signature

**DOI:** 10.1101/2025.01.30.25321355

**Authors:** David AA Baranger, Aaron J Gorelik, Margaret L Clapp Sullivan, Sarah E Paul, Alexander S Hatoum, Nico Dosenbach, Ryan Bogdan

## Abstract

**Background:** Task-based fMRI (tb-fMRI) provides experimentally controlled probes of brain function but has faced challenges in identifying reliable and behaviorally meaningful individual differences. Decoder models trained to distinguish task states - “neural signatures” - have been widely applied to characterize psychological processes, but their utility as individual-difference measures remains underexplored.

**Methods:** Using data from 9,024 early adolescents in the ABCD Study and 1,051 young adults in the Human Connectome Project, we derived a neural signature distinguishing high and low working memory loads in an Emotional N-back fMRI task, which captures individual differences in the whole-brain recruitment of working memory related neural resources.

**Results:** The signature robustly discriminated between task conditions in both samples (AUC=0.88-0.95). Signature expression was more reliable (ICC=0.43-0.53) and had larger associations with task performance, cognition, and psychopathology than standard estimates of regional brain activation in both adolescents and young adults (|**β**|=0.02-0.53). Signature performance reflected alignment between task-evoked activation and individual-differences effects. Relative to models trained to predict behavioral outcomes directly, the neural signature achieved comparable predictive performance while capturing unique variance and required substantially smaller training samples to achieve both directionally consistent out-of-sample associations (N=20 vs N=80-320) and maximal out-of-sample associations (N=320 vs N=2,560).

**Conclusions:** Task-state decoders (neural signatures) can serve as practical and interpretable summary measures of task-related brain function for individual-differences research. They generalize well across outcomes and samples without additional optimization and can be readily derived in extant data. Neural signatures provide a complementary strategy for quantifying behaviorally relevant variation in task-evoked activity.

## Introduction

A central goal in biological psychiatry is to identify neurobiological processes that account for individual differences in cognition, behavior, and risk for psychopathology^1^. Task-based fMRI (tb-fMRI) is uniquely positioned to probe such processes by experimentally manipulating cognitive and affective states. However, translating task-evoked activation into reliable and clinically meaningful individual-difference measures has proven challenging. Concerns regarding the reliability of regional activation measures^2^ and the small magnitude of brain–behavior associations^3^ have prompted critical re-evaluation of tb-fMRI approaches. These challenges have driven methodological advances, including the development of multivariate models^4^, use of large-scale datasets^5^, and adoption of longer scans^6^. These efforts highlight the need for approaches that retain the experimental specificity of tasks while improving measurement properties for individual-differences research.

One promising direction involves multivoxel pattern analysis (MVPA), multivariate decoder models also referred to as “neural signatures”. MVPA is a well-established class of approaches that estimate psychological or task states from distributed patterns of brain activity^7–10^. Such models have been extensively developed in domains including pain^11^, affect^12^, reward^13^, and craving^14^, where they have demonstrated sensitivity, specificity, and, in some cases, high reliability^15^. However, most work has focused on state-level decoding, and less is known about whether these models provide stable and behaviorally meaningful indices of individual differences^16–19^. This question is particularly relevant for biological psychiatry, where reliable neural phenotypes are needed to bridge experimental paradigms and clinical outcomes. If task-state decoders can provide stable summaries of how individuals recruit neural systems under cognitive demand, they may offer a powerful complementary approach to existing strategies for characterizing neurobiological variation associated with psychopathology.

Working memory (WM), the cognitive process in which information is stored and manipulated ^20^, is a complex higher order phenotype. It correlates with measures of executive function and cognitive ability ^20,21^, as well as with related cognitive constructs, including short-term memory^22^ and cognitive control^23^. Evidence of working memory deficits across psychiatric illnesses (e.g.,schizophrenia^24^, ADHD^25^, depression^26^, bipolar disorder^27^, substance use disorders^28^, and transdiagnostic measures of psychopathology^29,30^) have led to theories highlighting executive function deficits in the etiology of psychopathology^28,31,32^. WM-related brain activation is associated with broad and specific (e.g., cognitive control) measures of executive function and cognitive ability (e.g., IQ testing)^24^, and transdiagnostic psychopathology (e.g., schizophrenia, ADHD, depression, bipolar disorder, substance use disorders)^25–29^. Prior work has applied multivariate approaches to working memory and executive function^33,34^, including the role of sustained elevated activity^35^, the contribution of sensorimotor regions^36^, and identifying modality-independent processes^37^. MVPA analyses have additionally explored behavioral associations, finding that MVPA decoding of task load is correlated with behavioral performance^37–39^ and is sensitive to pain and threat manipulations^40,41^. However, these approaches have not been systematically evaluated as general-purpose individual-differences measures across large samples and diverse phenotypes.

Here, we evaluated whether a working memory task decoder can serve as a reliable and generalizable index of individual differences. We used data from the Emotional N-back fMRI task in the Adolescent Brain Cognitive Development^SM^ ABCD Study (ABCD Study®; data release 5.1), a large sample (N=11,875) of children and adolescents ages 9-12^42^. We test the accuracy and specificity of the signature’s predictions, as well as their replicability and stability. We then examine the association of the neural signature with cognition and mental health, comparing effect sizes to those observed in a traditional tb-fMRI and ML analysis. We leverage the large size of the ABCD study to explore the sample size requirements for the development of future neural signatures that are sensitive to individual differences. Finally, we test the generalizability of the neural signature in a fully independent sample, the Human Connectome Project (HCP), consisting of N=1,052 young adults (ages 23-35) who completed a visual working memory fMRI task.

## Methods and Materials

The ABCD study is a longitudinal study of late childhood and adolescence that has recruited over 11,000 participants from 21 sites across the United States. Participants were between the ages of 9-10 at the first study wave (referred to here as the ‘baseline’ wave). Neuroimaging data were collected every-other year, and data were retrieved (release 5.1) from the National Institute of Mental Health Data Archive (NDA; https://nda.nih.gov/), which included all available data from the baseline wave and the second follow-up wave (referred to here as ‘follow-up wave 2’).

### MRI Acquisition and Processing

Casey et al., 2018^42^ provide an in-depth description of the ABCD Study^®^ imaging acquisition protocol and parameters and Hagler et al., 2019^43^ provide an in-depth description of the ABCD Study^®^ image processing and analysis methods. The present analyses used tabulated neuroimaging data provided as part of the 5.1 data release. Participants completed two runs of an eight-block Emotional N-back working memory task^42^. Participants were presented with stimuli and asked to indicate whether or not they match a target image. In the low working-memory load ‘0-back’ condition, a target stimulus is presented at the beginning of each block, and participants are asked to indicate whether each subsequent stimulus matches the target image. In the high working-memory load ‘2-back’ condition, participants indicate whether a given stimulus matches the stimulus presented two screens prior. In the present analyses we used estimates of condition-level activation, averaged within ROIs defined by the Destrieux cortical atlas and FreeSurfer’s anatomically-defined subcortical ROIs^44–46^. See **Supplemental Methods** for additional details.

## Measures

### NIH Toolbox

The NIH toolbox is a cognitive battery consisting of seven different tasks that cover episodic memory, executive function, attention, working memory, processing speed, and language abilities and are used to generate three composite scores (**Supplemental Methods**)^47^.

### CBCL

The Child Behavior Checklist (CBCL)^48^ is a 113-item questionnaire on which caregivers rated items representing specific problems in the past six months (**Supplemental Methods**).

### Psychotic-like Experiences

Children completed the 21-item Prodromal Questionnaire-Brief Child Version (PQ-BC)^49^.

### Pubertal status

Parents and children both completed a 5-item scale on the child’s pubertal development^50^, combined to a summary score. The parent rating was used as the primary measure. The child rating was used if the parent rating was unavailable.

### Machine learning

#### Balanced split-halves

To facilitate machine learning training and testing without data leakage, the ABCD dataset was divided into two grouped splits, where all observations from the same site were assigned to the same fold, balanced on age, sex, number of visits with imaging data, pubertal status, in-scanner motion, and task performance (**Supplemental Table 1, Supplemental Methods**).

### Preprocessing

tb-fMRI activation estimates were processed with longCOMBAT to adjust for effects of imaging site on the data^51^. longCOMBAT preprocessing was performed separately for each of the two split-halves. Activation estimates were then standardized so that across both conditions each region had a mean of 0 and a standard deviation of 1.

#### Elastic-net

In each of the two balanced split-halves, a logistic elastic-net classifier^52^ was trained on brain activation data to distinguish between the 0-back and 2-back conditions (i.e., “is the brain map of activation from a given participant from the 0-back or 2-back condition?”; **Supplemental Methods**). In addition to the regression estimate of each feature, the encoding model was derived via the Haufe transformation as an interpretable and reliable feature importance metric (**Supplemental Methods**)^53,54^.

#### Out-of-sample performance

The final trained logistic elastic-net classifiers were used to predict condition probabilities in the held-out split (i.e., the model trained in Split 1 was tested in Split 2, and the model trained in Split 2 was tested in Split 1).

Individual difference measures

Linear elastic net regression models were trained using the identical split-halves on each individual difference outcome measure, using regional 0-back, 2-back, and 2-back>0-back activation measures.

### Reliability

Linear mixed effect models were used to assess the reliability (ICC 3,1) of the neural signature (i.e., predicted condition probabilities) and activation of each region^55^. Split-half reliability was computed using estimates derived from each of the two task runs. Longitudinal stability was computed using estimates from each wave. Analyses were further restricted to only include one member from each family, as the collinearity between family and subject identifiers can downwardly bias reliability estimates. Reliability analyses included a random intercept for site, and fixed effects for split, mean in-scanner motion, and scanner manufacturer.

### Individual differences

Linear mixed-effect models^56,57^ were used to test the association between the 2-back>0-back neural signature, 2-back>0-back regional activation, and linear elastic net model regression predictions with individual difference measures, including task performance, NIH toolbox performance, and psychopathology (CBCL and PQ- BC). All variables were winsorized (+/- 3SDs) prior to analyses to reduce the influence of outlier observations, measures of psychopathology were log-transformed due to high skew, and all variables were z-scored prior to analyses so that effect sizes would be comparable across outcomes. Models included random intercepts for participant, family, and site. Fixed effects included split, age, age^2^, sex, pubertal status, familial relationship (i.e., sib, twin, triplet), average in-scanner motion, and scanner manufacturer. Results were false discovery rate (fdr) corrected for multiple comparisons. Follow-up analyses additionally adjusted for: 1) task performance (0-back and 2-back accuracy) and 2) additional demographic variables, including parental income and education, and participant race and ethnicity. Whole-brain maps (i.e., activation, feature importance, or associations with cognition/psychopathology) were compared using a pairwise correlation, with p-values that are robust to spatial-autocorrelation (p_robust_) computed via permutation (**Supplemental Methods**).

### Bootstrapped analyses

Bootstrapped analyses examined the influence of training sample size on model performance and neural signature associations. Baseline wave data from unrelated participants in Split 1 was resampled with replacement, maintaining an equal representation of each of the 10 sites. Neural signature and machine learning models were trained, and then were tested in baseline wave data from unrelated participants in Split 2. Machine learning models were trained to predict 2-back accuracy as ML models trained on accuracy achieved cross-trait predictions comparable to the neural signature. The sample size of the Split 2 testing data was held constant throughout.

### Generalizability

Generalizability analyses used the S1200 2017 release of the Human Connectome Project (HCP)^58,59^, a family-based study (2-4 siblings per family, most including a twin pair) of 1206 healthy young adult participants (ages 23-37). The final analytic subsample included N=1,051 participants from n=439 families. Analyses used the provided processed N-back task fMRI data (**Supplemental Methods**)^60,61^.

### Measures

#### Cognition

As in the ABCD, participants in the HCP completed the NIH toolbox^47^. Participants additionally completed measures of sustained attention, verbal episodic memory, spatial orientation, and fluid intelligence (**Supplemental Methods**)^62,63^.

#### ASR

The Achenbach Self-Rerport (ASR)^64^ is a 123-item questionnaire on which participants rated items representing specific problems. These were used to generate syndrome and DSM-oriented scales, as well as composite scales.

#### Analyses

N=41 participants completed a second MRI scan within one year (18-328 days, mean= 136, sd= 65). Linear mixed effect models were used to assess the reliability (ICC 3,1) of the neural signature (i.e., predicted condition probabilities) and activation of each region^55^. Reliability analyses further included fixed effects for mean in-scanner motion. Linear mixed-effect models^56,57^ were used in the full HCP sample to test the association between the neural signature (trained in ABCD) and regional activation with individual difference measures. All variables were winsorized (+/- 3SDs) prior to analyses to reduce the influence of outlier observations, and measures of psychopathology were log-transformed due to high skew. All variables were z-scored prior to analyses so that effect sizes would be comparable across outcomes. Models included random intercepts for family and fixed effects for age, age^2^, sex, familial relationship (i.e., MZ-twin, DZ-twin, sibling, half-sibling), and average in-scanner motion. Results were false discovery rate (fdr) corrected for multiple comparisons.

## Results

The ABCD Study sample comprised 13,477 observations from 9,024 participants who had usable Emotional N-back tb-fMRI data for at least one wave. As previously reported^5,65^, the N-back task activated regions across the bilateral frontoparietal and salience networks, as well as the bilateral thalamus and caudate (**Figure 1a**).

**Figure 1.**
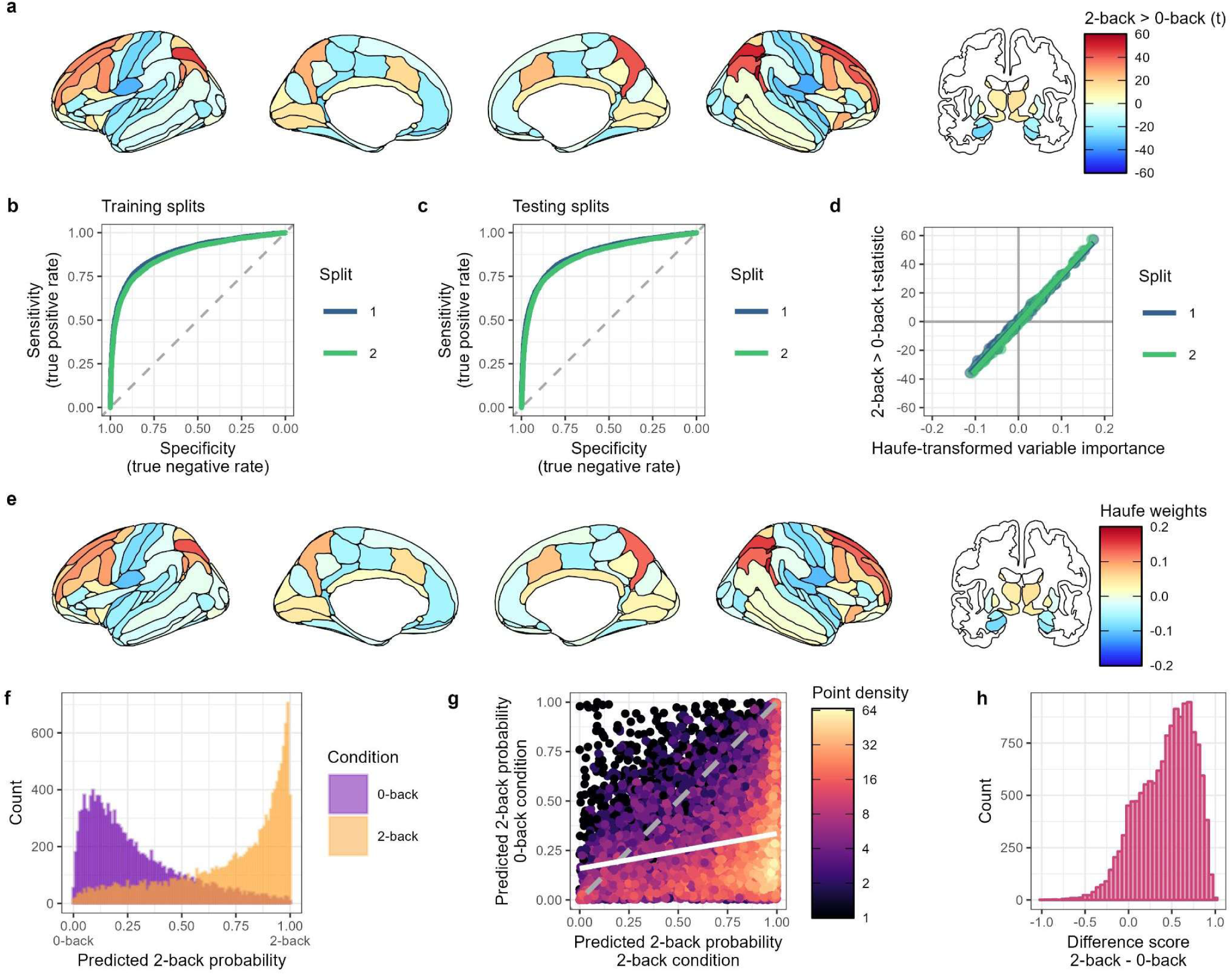
Emotional n-back neural signature. **A**) Main-effect of the 2-back > 0-back contrast in the N-back task, using 13,477 observations from 9,024 participants. Colors reflect the t-statistic of the test. **B&C**) An elastic-net classifier distinguishing high and low working memory conditions was trained. Model performance - area under the receiver operating characteristic curve (AUC) - was comparable across balanced split-halves in both the training splits (**B;** Split 1 AUC=0.884; Split 2 AUC=0.872) and testing splits (**C;** Split 1 AUC=0.877; Split 2 AUC= 0.867). **D)** A Haufe-transformation of variable (brain region) weights identified the contribution of individual regions to model performance. Plot shows the correlation of the haufe-transformed variable importance with the main-effect of the task in both splits (Split 1: r=0.99, p_robust_=7.5x10^-14^; Split 2: r=0.99, p_robust_=1.0x10^-13^). Points are individual regions. **E**) Visualization of the haufe-transformed variable importance of each brain region, averaged across splits. Variables with larger absolute weights contribute more to the model’s predictions. **F)** Histogram of model predictions for each MRI scan for each of the two conditions (purple: 0-back; orange; 2-back), combined across data splits. Values reflect the predicted probability that a scan is from the 2-back condition, given a forced choice between 2-back and 0-back. 1.00 = 100% probability that a scan is 2-back; 0.00 = 0% likely it is 2-back and thus 100% probability it is 0-back; 0.5 = both conditions are equally likely. **G)** Model predictions for the two conditions for each individual scan (points), colored by density (number of overlapping points). The white line reflects the correlation between predictions for the two conditions. **H)** Histogram of the difference between the model predictions for the two conditions. Values reflect how separable the two conditions are for each scan. Positive values indicate that the 2-back condition had a higher 2-back probability than the 0-back condition .

### A neural signature for the emotional working memory task

To generate a neural signature of the Emotional N-back task, a logistic elastic-net classifier was trained on brain activation data. Performance of the elastic-net classifier was comparable (**Figure 1b&c**) across training (Split 1 AUC=0.884; Split 2 AUC=0.872; AUC=area under the curve) and testing (Split 1 AUC=0.877; Split 2 AUC= 0.867) rounds, indicating minimal overfitting. Haufe-transformed variable weights (**Figure 1e**) were strongly correlated with the main effect of task on regional activation in both splits (Split 1: r=0.99, p_robust_=7.5x10^-14^; Split 2: r=0.99, p_robust_=1.0x10^-13^; **Figure 1d)**, and was similar across splits (r=0.99, p_robust_=7.5x10^-14^ **Supplemental Figure 1f**). Model predictions for the 0-back and 2-back conditions (**Figure 1F**) reflect the predicted probability that a scan is from the 2-back condition, given a forced choice between the two conditions. Model predictions were moderately correlated across the two conditions (r=0.23; **Figure 1g**), much lower than the correlation in activation between conditions in individual regions (rs=0.53 - 0.85; **Supplemental Figure 2**). A difference score between the predictions for the two conditions was computed (**Figure 1h**), to reflect how working memory load modulates the signature response within-subject. *Post-hoc* supplemental analyses using 6 other linear and non-linear machine learning algorithms found that all performed comparably, had identical haufe-transformed variable weights, and similar out-of-sample predictions (**Supplemental Methods**, **Supplemental Figure 3**).

### Improved reliability and sensitivity to individual differences

The reliability and stability of the difference score (ICC=0.43-0.47; **Figure 2**; **Supplemental Table 2**) was significantly greater (p<0.05 bonferroni-corrected) than every individual region and resulted in an 8-15-fold improvement over the median regional reliability for the 2-back>0-back condition, and a 2-3x improvement over the maximum regional reliability (**Supplemental Methods & Data**). The 0- and 2-back predictions were similarly significantly more reliable and stable (p<0.05 bonferroni-corrected) than the majority of regions (split-half reliability: 83-98% of regions; longitudinal stability: 76-96% of regions).

**Figure 2.**
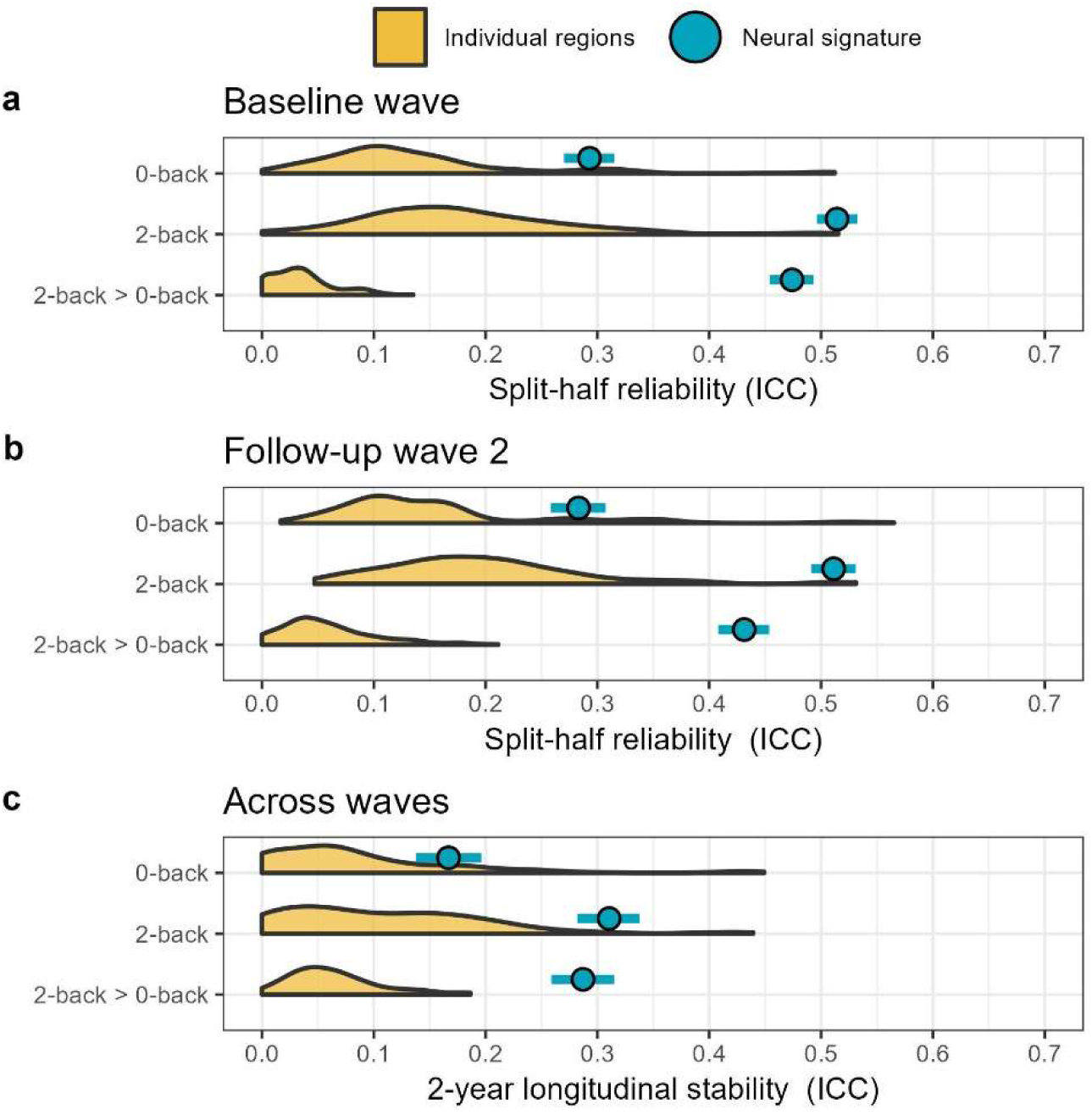
Reliability of the Neural Signature and Individual Regional Activation. The split-half reliability (comparing runs within one scan) and longitudinal stability (comparing two scans) was examined for neural signature predictions during the 0-back condition, 2-back condition, and their difference (2-back > 0-back). Blue points are the neural signatures, with 95% confidence intervals indicated by horizontal lines. ICC=intraclass correlation coefficient. Shaded yellow regions represent the distribution of reliabilities across all regions (n=167) for each of the corresponding conditions and contrasts. Analyses were restricted to unrelated participants. **A)** Split-half reliability in the baseline session, N=6,551. **B)** Split-half reliability in the year 2 follow up, N=5,386. **C)** Longitudinal stability across the two waves, N=4,111.

The signature was significantly associated with all measures examined (p<0.05 false discovery rate (fdr) corrected). Associations with in-scanner behavioral performance (e.g., 2-back accuracy, ꞵ=0.34; **Fig. 3a**) and neuropsychological assessments of cognition (e.g., total cognitive composite score, ꞵ=0.31; working memory performance, ꞵ=0.24; **Fig. 3b**) were positively associated with the neural signature. Indices of psychopathology (e.g., social problems, ꞵ=-0.079; attention problems, ꞵ=-0.076; psychotic-like experiences, ꞵ=-0.064; **Fig. 3c**) had smaller associations with the neural signature. Effects exceeded those of individual regions for almost all outcomes (**Supplemental Data**) and were robust to additional demographic and task-performance covariates (**Supplemental Data**). Across all outcomes, the strength and direction of the neural signature association corresponded to the alignment between the regional association map and the main-effect-of-task map (**Supplemental Figures 4-6**). Larger neural signature associations were also observed for outcomes with more significant individual regional associations (**Supplemental Figure 4**). *Post-hoc* analyses revealed that neural signature associations are unlikely to be attributable to uncontrolled motion confounding or overall task engagement (**Supplemental Results**) and no evidence of sex moderation was observed (**Supplemental Data**).

**Figure 3.**
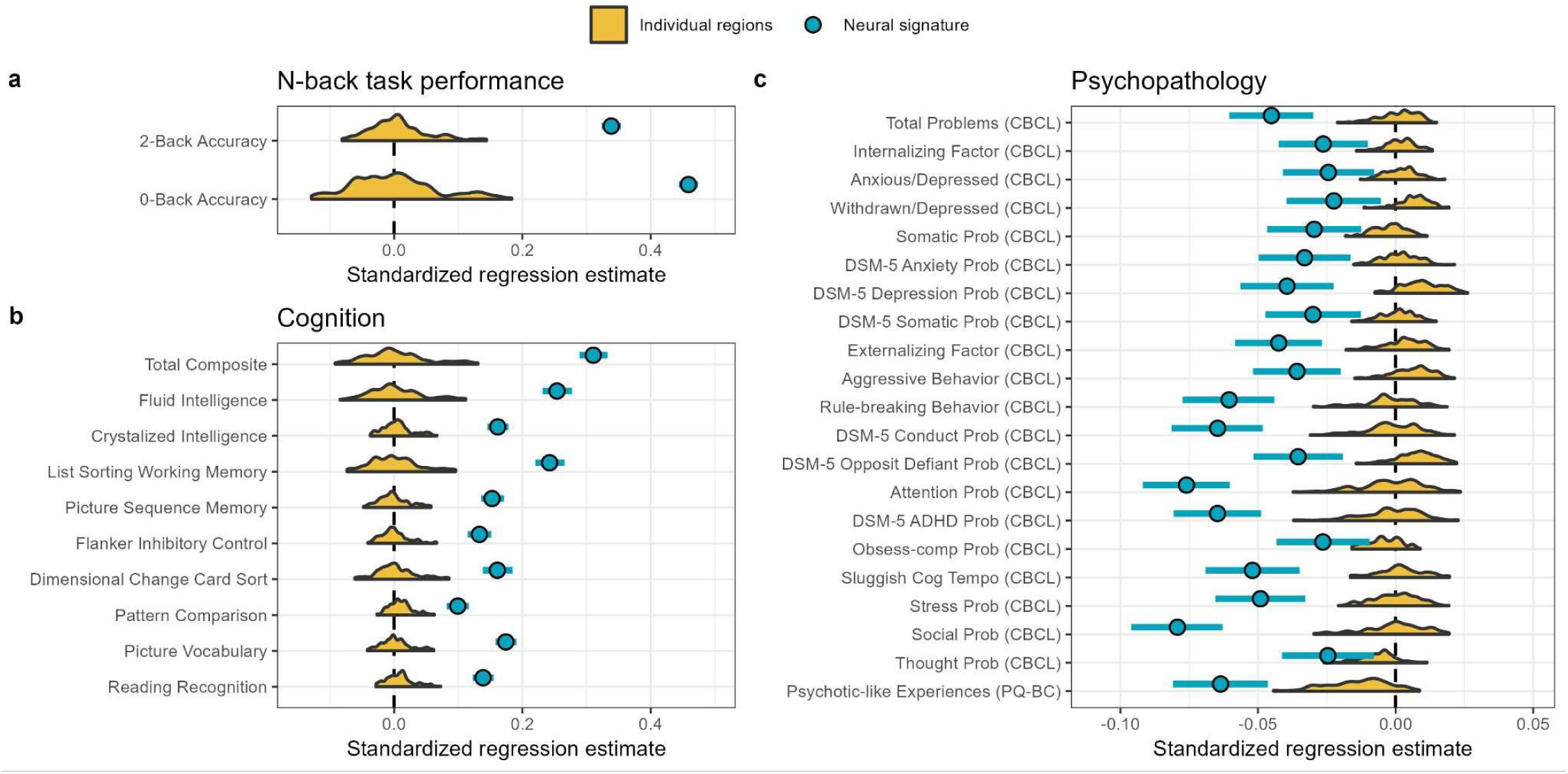
The neural signature is associated with individual differences in behavior, cognition, and psychopathology. The neural signature: positively correlates with **A**) N-back task performance, **B**) positively correlates with NIH toolbox task performance and composite measures, and **C**) negatively correlates with psychopathology. Shaded yellow represents the distribution of effects across all regions. Blue points reflect neural signature association estimates with 95% Confidence intervals (horizontal lines). Estimates reflect standardized regression coefficients. Note: Directionality of neural signature associations (e.g., positive for cognition and negative for psychopathology) are examined in the section *Neural signature associations reflect correspondence with the main effect of task*.

### Comparison with other multivariate approaches

Machine learning (ML) models trained to predict each individual outcome achieved similar performance to the neural signature (**Figure 4a & 4c**), though in contrast to the neural signature, associations for 11 of the 21 psychopathology outcomes were non-significant (**Supplemental Data**). The model fit (Akaike information criterion, AIC) improvement significantly differed (p-fdr<0.05) between the ML and neural signature models for thirteen outcomes (**Supplemental Methods**), where-in the ML model was largely a stronger predictor of cognition, and the neural signature was largely a better predictor of psychopathology. When the ML model and neural signature predictions were simultaneously entered into regressions, the overall direction of effect remained consistent with models that included each separately, and both had significant unique associations (**Figure 4D**). Analyses further tested the association of each ML model with every other outcome. The neural signature showed comparable generalizability to the top-performing ML models—the models trained to predict 2-back accuracy (**Figure 4b**) and the Total Composite score (**Supplemental Figure 7**).

**Figure 4.**
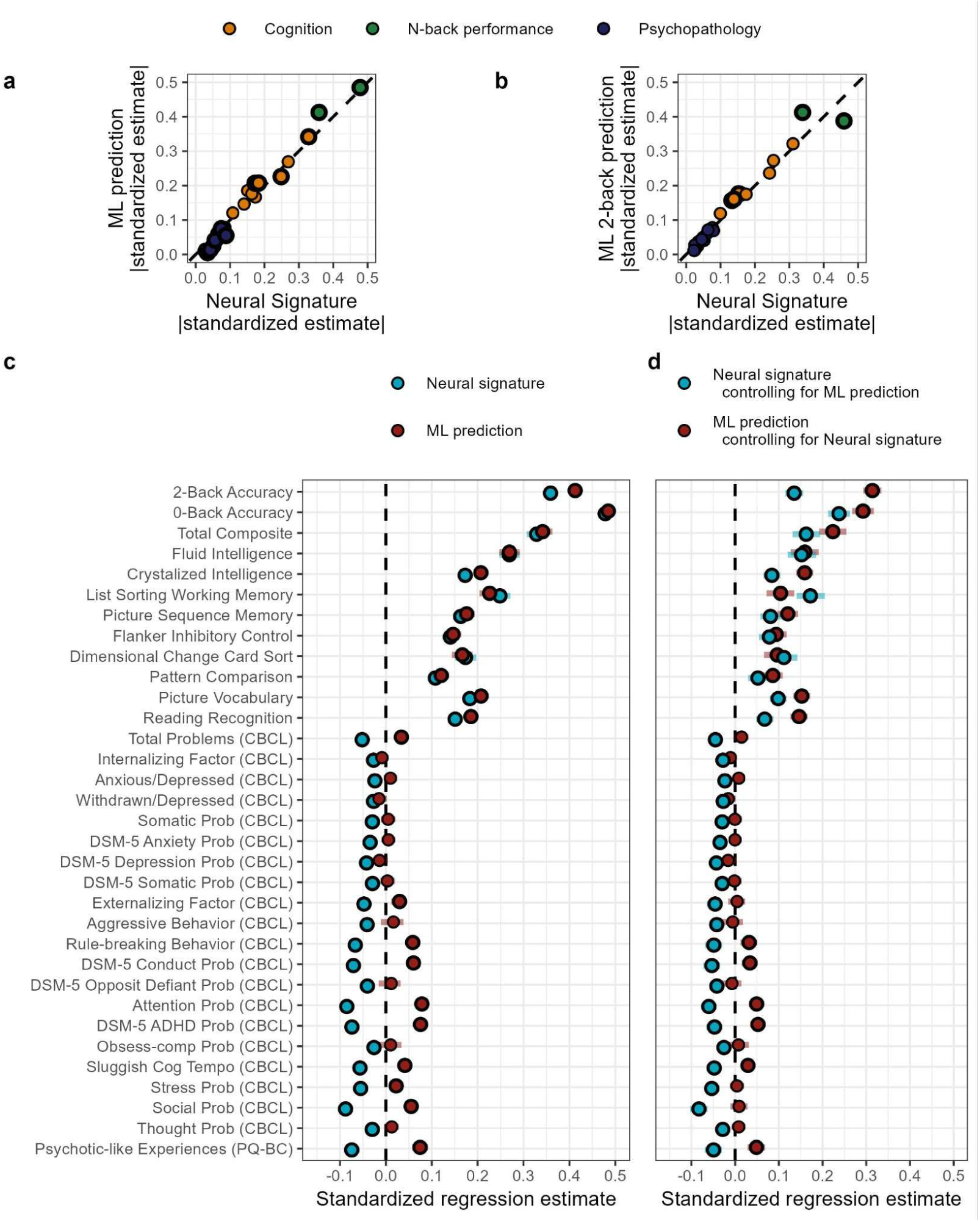
The neural signature achieves similar performance to machine learning models. **A)** The absolute value of the standardized regression estimate from models predicting each outcome using the neural signature or a linear elastic net regression machine learning (ML) algorithm. The dashed line indicates the diagonal. Colors reflect the type of individual difference measure. Bolded points indicate the model fit statistics for the neural signature and ML models significantly differ from each other after false discovery rate correction for multiple comparisons at p<0.05. **B)** The absolute value of the standardized regression estimate from models predicting 2-back accuracy. **C)** Standardized regression estimates from models that included either the neural signature or predictions from a linear elastic net regression machine learning (ML) algorithm. Lines reflect the 95% confidence intervals. Bolded points survive false discovery rate correction for multiple comparisons at p<0.05. Note: ML model associations are largely positive, as models were separately trained to predict each outcome. Neural signature associations (e..g, positive for cognition and negative for psychopathology) are the same as presented in **Figure 3**. **D)** Standardized regression estimates from models that included both the neural signature and predictions from a linear elastic net regression machine learning (ML) algorithm.

### The impact of training sample size

Bootstrapped analyses examined the influence of training sample size on model performance and associations with behavior (**Figure 5**). For the neural signature, there were marked improvements in all performance measures from N=80 to N=320, at which point improvements began to taper. ML models attained comparable effect sizes to neural signature models at larger sample sizes, but performed substantially worse at lower sample sizes, yielding associations with individual difference measures near 0 or with the opposite sign at samples less than N=80-160. Simulation analyses with synthetic data (**Supplemental Methods**) confirmed that signature and linear regression prediction models largely attain comparable performance, but that signature models achieved greater performance at smaller training samples, particularly when effect sizes were small (**Supplemental Figures 8-10**). Power analyses estimated the total sample size (training + test) needed to detect each association with 80% power, finding that the sample size needed by the neural signature model was, on average, 50-66% smaller than the sample size needed by the ML model (**Figure 5d**).

**Figure 5.**
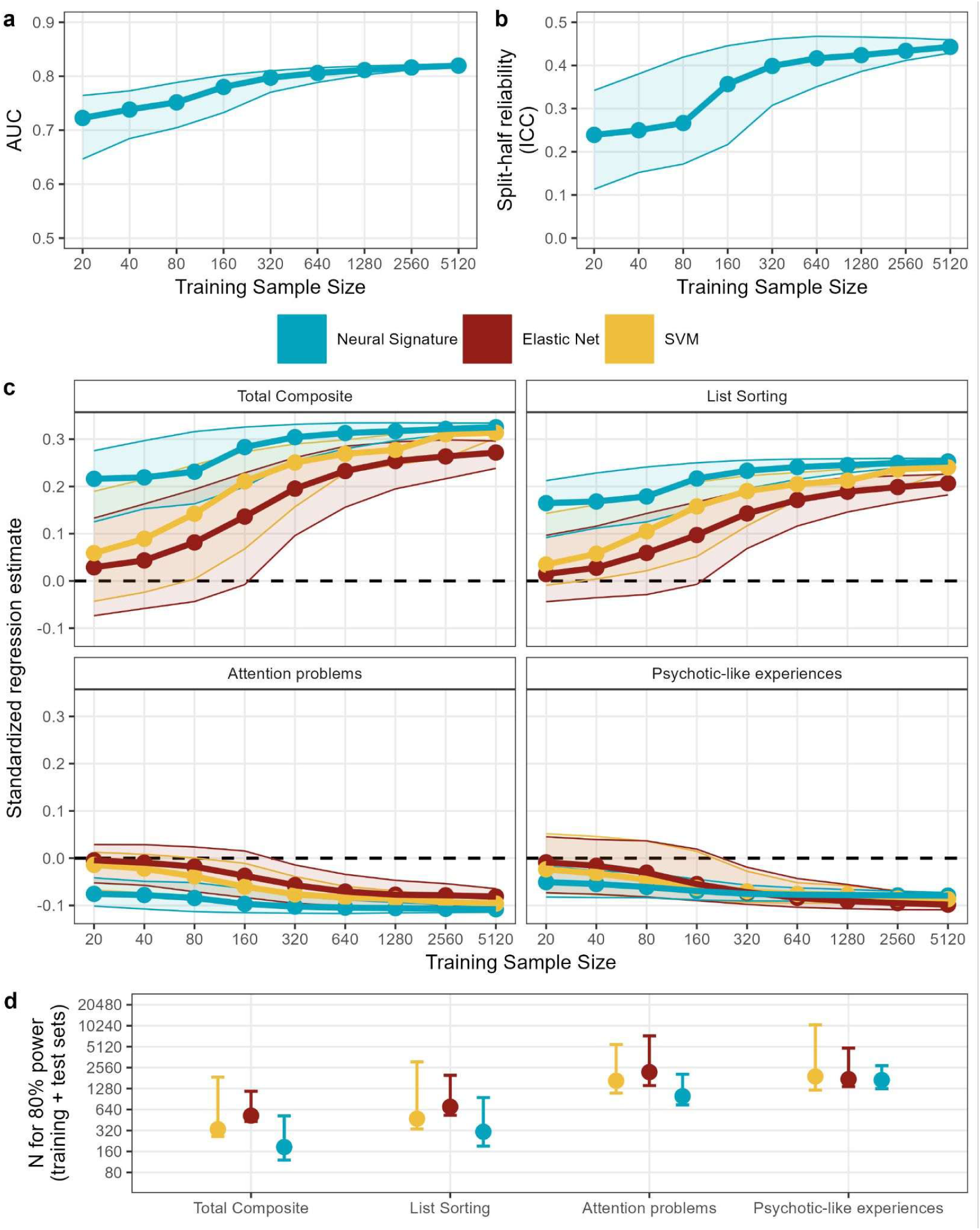
Influence of training sample size. Bootstrapped models were trained on samples from N=20 to N=5,120, repeated 2,000 times per sample size. Training and testing samples were restricted to the baseline visit. Points reflect the median across bootstraps, and shaded areas the 95% confidence interval. **A)** The area under the curve (AUC; Y axis) of model predictions. **B)** The split-half reliability (ICC - intraclass correlation coefficient) of the 2-back>0-back signature. **C)** Standardized regression estimates from multilevel models testing the association of the signature (blue) and machine learning model predictions ( trained to predict 2-back accuracy; red: Elastic Net; yellow: SVM) with key individual difference measures - the Total Composite and List Sorting working memory scores, Attention problems, and Psychotic-like experiences. **D)** Using the results in ‘C’, power analyses estimated the sample size needed to attain 80% power at each training sample size. The minimum total size (training + test) was then found (points). Confidence intervals reflect the total sample size for the corresponding confidence intervals in ‘C’.

### Replication in an independent sample of young adults

To assess the generalizability of the signature, analyses tested its performance in the Human Connectome Project (HCP)^58,59,61^. The whole-brain pattern of the main effect of task load (2-back > 0-back) was similar to that observed in ABCD (r=0.93, p_robust_=5.8x10^-6^), though the HCP showed greater activation and de-activation across all regions (**Figure 6a**). The neural signature trained in the ABCD was then tested in the HCP sample. As in the ABCD sample, the signature classified task conditions (AUC = 0.95; **Figure 6b&c**) and attenuated the correlation between conditions (**Figure 6d**), yielding an individual difference measure with sufficient variability (**Figure 6e**). The reliability of the neural signature was comparable to the reliability observed in ABCD (**Figure 6f**, ICC = 0.53). Individual difference analyses (**Figure 6G-I, Supplemental Data**) found significant associations with cognition, behavior, and mental health for all phenotypes but 5 following FDR correction (**Figure 6g-i, Supplemental Data**). As in the ABCD Study, the signature showed stronger associations with task performance and behavioral tasks than single regions (**Supplemental Data**). Analyses examined the influence of parcellation resolution, finding only a very modest effect on AUC (0.95-0.975) and no effect on individual difference associations (**Supplemental Figure 11**).

**Figure 6.**
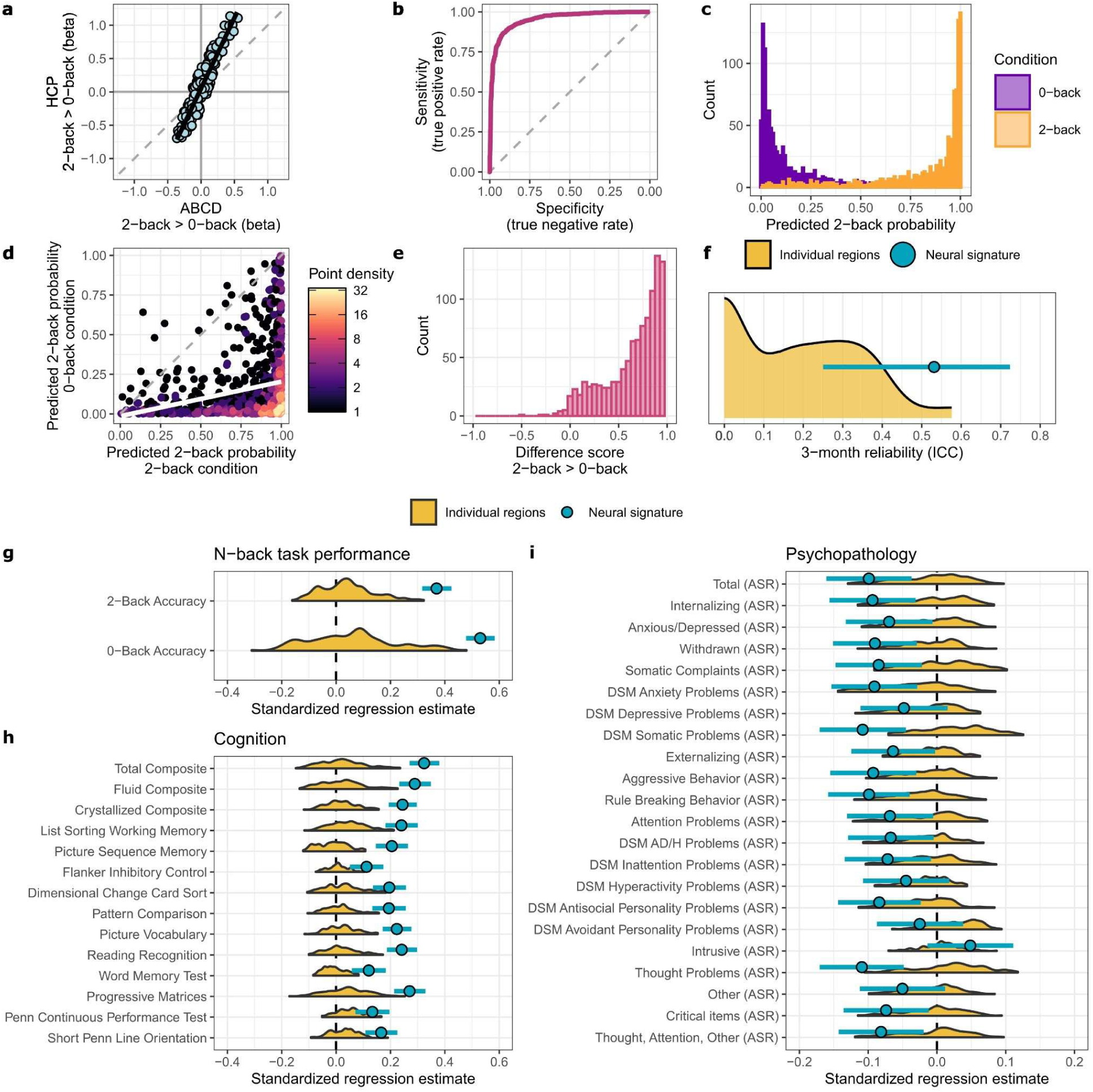
Replication in the Human Connectome Project. The neural signature trained in the ABCD was tested in the independent Human Connectome Project (HCP; N = 1,051)). **A)** Similarity of the main-effect of task between the ABCD and HCP samples (r=0.93, p_robust_=5.8x10^-6^). **B)** Receiver operating curve of task condition classification (AUC = 0.95). **C)** Histogram of model predictions for each MRI scan for each of the two conditions (purple: 0-back; orange; 2-back). **D)** Model predictions for the two conditions for each individual scan (points), colored by density (number of overlapping points). The white line reflects the correlation between predictions for the two conditions. **E)** Histogram of the difference between the model predictions for the two conditions. **F)** 3-month reliability of the 2-back>0-back contrast and neural signature in N=42 participants. Blue points is the neural signature, with 95% confidence intervals indicated by horizontal lines. ICC=intraclass correlation coefficient. Shaded yellow regions represent the distribution of reliabilities across all regions (n=167). **G-H)** Associations of the neural signature with individual difference measures, including N-back task performance, cognitive task performance and composite measures, and psychopathology. Shaded yellow represents the distribution of effects across all regions. Blue points reflect neural signature association estimates with 95% Confidence intervals (horizontal lines). Estimates reflect standardized regression coefficients.

## Discussion

We developed a neural signature of the Emotional N-back working memory task in a sample of adolescents (**Figure 1**) that is characterized by higher reliability and longitudinal stability (**Figure 2**), as well as larger associations with behavioral and mental health phenotypes (**Figure 3**) than traditional indices of working memory neural activity. The signature and these findings generalize to an independent sample of young adults from the Human Connectome Project (**Figure 6**). We further demonstrate that the neural signature approach performs well in settings where current standard machine learning approaches struggle. The neural signature approach requires less training data than standard ML approaches (**Figure 5**) and does not require external phenotypes, while still yielding relevant insights into how global patterns of activation differ between individuals. Overall, the tb-fMRI neural signature approach has potential to further our understanding of the neurobiological correlates of cognition and behavior and enhance the use of tb-fMRI data acquired in existing studies as well as its implementation in future research.

The neural signature can be conceptualized as a latent factor or generative model that separates conditions^53^. Under this view, we would expect the signature to reflect the contribution of key working memory regions, which is precisely what we observed - feature weights showed that the signature reflects activation of regions in the fronto-parietal and salience networks, as well as the bilateral caudate and thalamus that have been implicated in working memory^66,67^. Indeed, the feature-importance map of the signature recapitulated the univariate association map (rs=0.99; **Figure 1**) and signature associations reflect correspondence between the main-effect of task map and the regional-association map (**Supplemental Figure 4**). This suggests that the signature indexes a dimension of coordinated brain activity, characterized by activation of frontoparietal systems and deactivation of default-mode regions, that varies across individuals and is relevant for both cognitive performance and psychopathology. Positive associations with cognition indicate that participants with higher scores on cognitive tasks exhibit relatively greater activation of task-positive regions (e.g., fronto-parietal and salience networks), as well as relatively greater de-activation of task-negative regions (e.g., default-mode network, motor and visual networks). In contrast, negative associations with psychopathology reflect that participants with worse mental health exhibit less activation of task-positive regions and less de-activation of task-negative regions.

These observations align with and extend prior work finding that task performance correlates with the divergence of activation between load conditions during working memory^68^, and that regions showing the strongest effect of WM load are also the most strongly associated with WM task performance^69^. The broad associations with the signature further align with prior work showing that working memory is associated with a wide range of cognitive and behavioral measures^22,70,71^, reflecting its central role in cognition. Associations with childhood and adult psychopathology were strongest for psychotic-like experiences and externalizing symptoms, including attention problems and ADHD, and were broadly consistent across developmental stages. Associations in these domains with regional WM activation are well-documented in adults^72–76^, but the extent to which children with these symptoms displayed similar patterns of activation has remained an open question^29,77–79^. Our findings also bring clarity to the question of whether WM neural activation disruption is specific to a subset of psychopathologies^29^. While the strength varied across domains, nearly all measures examined were associated with the signature. Further, models of psychopathology frequently point to the deactivation of frontoparietal areas, particularly the dorsolateral prefrontal cortex, as a key mechanism by which working memory influences risk^5,65,80–82^. The current analyses extend this work by demonstrating that these associations reflect a whole-brain pattern of activation, rather than effects localized to specific regions, suggesting that psychopathology is associated with global differences in how neural resources are recruited during working memory.

Traditional contrast-based tb-fMRI estimates are limited by shared variance across conditions^83^. By explicitly optimizing discrimination between conditions, the neural signature reduces this shared variance, yielding a more reliable individual-differences measure. While the reliability attained can be described as ‘fair’^84^ (ICCs=0.43 - 0.53), it yields a 2x - 8x reduction in the required sample size for individual difference studies, relative to the reliability of regional activation in the ABCD and HCP studies (**Supplemental Fig 12)**. Our findings thus converge with prior work on multivariate models, including both machine learning (ML) and factor-analysis, which has found that multivariate models frequently yield larger effect sizes^85–90^ and more reliable predictions^15,91^ than single regions, as the model can make use of variance that is shared across features to reduce measurement error^87^. Interestingly, while the neural signature achieved comparable performance to standard ML models, analyses with both model predictions jointly entered showed that the neural signature captures variance that is partially unique from standard ML approaches (**Figure 4**). This may reflect a bias-variance tradeoff, wherein the signature is more sensitive to individual differences that occur along the latent axis defined by the two task states, whereas a more flexible ML approach will better detect associations outside that latent axis for the specific outcome phenotype. Notably, while the neural signature and a standard ML approach performed comparably at larger sample sizes, the signature achieved greater performance at smaller sample sizes, and required a smaller sample size to be powered to detect associations.

The neural signature approach is not without limitations. Simulations emphasized that signature performance is dependent on accurate condition classification - if task conditions are too similar the signature approach would not be expected to provide any benefits (**Supplemental Figure 9**). Simulations also showed that the neural signature is effective because the regions with the strongest individual difference associations are the ones that are the most differentially de/activated (**Supplemental Figure 10**)^69^. As such, it would not be expected to yield enhanced associations if individual difference effects are highly localized, or in tasks that produce large activations in off-target systems (**Supplemental Figures 13-15**). That is, working memory may represent a favorable context for this approach. Working memory tasks produce robust, spatially distributed activation patterns that align closely with behavioral performance, which may facilitate the construction of reliable signatures. While the reliability of the neural signature was substantially greater than that of any individual region, it was overall much lower than common structural measures from MRI^2^. This places limitations on the magnitude of effect size that can be detected and precludes many tests of interaction effects^92^. Further improvements are needed for tb-fMRI to achieve clinical relevance on an individual level. Recent evidence suggests that hierarchical models which pool information across trials yield considerably improved activation reliability^93^, with preliminary evidence that they also improve signature predictions^94^. Limitations of the current analyses also include that the mental health measures we used from the ABCD study rely on parent-report, which may be biased^95^. Further, some data suggest that externally observable behavior, such as externalizing, is more likely to be captured by parent-report as opposed to potentially unobservable internalizing behavior^96,97^.

The low reliability and small effects of traditional tb-fMRI research places significant constraints on its utility. Here, we show that a neural signature approach results in significantly higher reliability and larger effects, as well as insights into the neurobiological correlates of individual difference measures. The neural signature approach brings the strengths of multivariate modeling to topic areas where large samples may be difficult or too costly to acquire, including when the prevalence or characteristics of a population hinder the collection of large samples^98,99^, and when the task-paradigm of interest is less widely adopted. Further, as outcome measures of interest do not need to be present for model training, this approach can facilitate study designs wherein a signature is developed in a larger, but less well-phenotyped sample, and then associations in smaller, but more deeply-phenotyped samples are examined. The neural signature approach can also boost reliability in existing data sets, thereby improving the return on the millions of dollars already invested into task fMRI. Neural signatures show promise as complementary individual difference analysis for task fMRI.

## Acknowledgements

This study was supported by K99AA030808 (DAAB) and R01DA54750 (RB). Additional funding included: AJG (DGE-213989), SEP (F31AA029934), ASH (K01AA030083), RB (R21AA027827, U01DA055367). Data for this study were provided by the Adolescent Brain Cognitive Development (ABCD) study which was funded by awards U01DA041022, U01DA041025, U01DA041028, U01DA041048, U01DA041089, U01DA041093, U01DA041106, U01DA041117, U01DA041120, U01DA041134, U01DA041148, U01DA041156, U01DA041174, U24DA041123, and U24DA041147 from the NIH and additional federal partners (https://abcdstudy.org/federal-partners.html). A listing of participating sites and a complete listing of the study investigators can be found at https://abcdstudy.org/consortium_members/. ABCD consortium investigators designed and implemented the study and/or provided data but did not necessarily participate in the analysis or writing of this report. This manuscript reflects the views of the authors and may not reflect the opinions or views of the NIH or ABCD consortium investigators. Data were also provided in part by the Human Connectome Project, WU-Minn Consortium (Principal Investigators: David Van Essen and Kamil Ugurbil; 1U54MH091657) funded by the 16 NIH Institutes and Centers that support the NIH Blueprint for Neuroscience Research; and by the McDonnell Center for Systems Neuroscience at Washington University. Computations were performed in part using the facilities of the Washington University Research Computing and Informatics Facility (RCIF). The RCIF has received funding from NIH S10 program grants: 1S10OD025200-01A1 and 1S10OD030477-01.

## Data availability

Data used in the preparation of this article were obtained from the Adolescent Brain Cognitive Development (ABCD) Study (https://abcdstudy.org), held in the NIMH Data Archive (NDA). This is a multisite, longitudinal study designed to recruit more than 10,000 children aged 9–10 and follow them over 10 years into early adulthood. The ABCD data repository grows and changes over time. The ABCD data used in this report came from the 5.1 release, available at https://nda.nih.gov/study.html?id=2313. Data additionally came from the Human Connectome Project (HCP) Young Adult, which includes imaging data on over 1100 healthy young adults, available at https://www.humanconnectome.org/. Results from this study, including the neural signature model weights, are available in the Supplement. Code for applying these weights to new data is available in the associated github repository: https://github.com/WashU-BG/NeuralSignature.

## Code availability

Analysis code is available at https://github.com/WashU-BG/NeuralSignature.

