## Supplement for "Task-State Decoders as Individual-Differences Measures: Evidence from a Working Memory Neural Signature"

**Contents:**

Supplemental Methods: 3 - 8

Supplemental Results: 9

Supplemental References: 10-11

Supplemental Tables 1-3: 12 - 14

Supplemental Figures 1-15: 15 - 29

**Supplemental Methods**

ABCD IRB: All ABCD study procedures were approved by a central Institutional Review Board (IRB) at the University of California, San Diego, and by individual site IRBs (e.g. Washington University in St. Louis, protocol # 160091). Parents or guardians provided written informed consent after the procedures had been fully explained and children assented before participation in the study. Use of ABCD data for secondary data analysis was also approved by the Washington University in St. Louis IRB (protocol # 201708123).

ABCD MRI data acquisition and processing

Structural MRI: 1 mm isotropic T1-weighted (T1w) structural magnetic resonance images (MRI) were acquired on 3 Tesla (Siemens, Phillips and GE) MRI scanners using either a 32-channel head or 64-channel head-and-neck coil. Scan protocols were carefully harmonized across the three MRI vendor platforms to reduce scanner-caused variability. MRI data were processed with the Multi-Modal Processing Stream software package that includes FreeSurfer 5.3. Further processing included a modified intensity normalization process. A description of the quality-control measures conducted on the processed data is provided in Hagler et al. MRI analyses included only participants whose structural MRI reconstructions passed QC tests.

Functional MRI: Participants completed two runs of an eight-block Emotional N-back working memory task[1]. Participants were presented with stimuli and asked to indicate whether or not they match a target image. In the low working-memory load '0-back' condition, a target stimulus is presented at the beginning of each block, and participants are asked to indicate whether each subsequent stimulus matches the target image. In the high working-memory load '2-back' condition, participants indicate whether a given stimulus matches the stimulus presented two screens prior. The task included two runs, each with four 0-back and four 2-back blocks, with 10 2.5 second trials per block, and four 15 second fixation blocks[1]. A second dimension of the Emotional N-back is that stimuli varied with regard to their emotional content. Half of all blocks used images of places, while the other half used images of faces, with either happy, fearful, or neutral expressions. Behavioral measures include accuracy and reaction time in the 0-back and 2-back conditions.

2.4 mm isotropic BOLD (blood-oxygen level dependent) functional magnetic resonance images (fMRI) were acquired on 3 T (Siemens, Phillips and GE) MRI scanners. Scans used multiband EPI with a slice acceleration factor of 6 (TR=800ms, TE=30ms, Flip Angle=52°). Preprocessing included corrections for head motion, $B_0$ distortion, gradient nonlinearities, and between-scan motion[2–5]. fMRI images were then registered to T1w images, initial frames were removed, and voxel time series were normalized and sampled onto the cortical surface. Average ROI time courses were calculated for cortical surface-based ROIs using FreeSurfer's anatomically-defined parcellations and subcortical ROIs[6–8]. In the present analyses we use estimates from the Destrieux cortical atlas[9]. Time points with an FD greater than 0.9 mm were censored and time courses were temporally filtered using an infinite impulse response (IIR) notch filter to attenuate signals in the range of 0.31–0.43 Hz[10]. Effects of task on activation were computed using a general linear model[4]. Regressors included baseline and quadratic trends in the time-series data, as well as motion estimates and their derivatives[2]. Hemodynamic response functions were modeled as square waves convolved with a two parameter gamma basis function[2]. Task models include stimulus timing for each condition and linear contrasts of conditions.

ABCD Measures

NIH Toolbox: The NIH toolbox is a cognitive battery consisting of seven different tasks that cover episodic memory, executive function, attention, working memory, processing speed, and language abilities and are used to generate three composite scores[11]. Tasks include: 1) picture vocabulary (language and verbal intellect), 2) reading (language), 3) pattern matching (processing speed), 4) list sorting (working memory), 5) memory (episodic memory), 6)response inhibition (attention and executive function), and, 6) card sort (executive function and cognitive flexibility). Higher order factors including a Total Composite (all seven tasks), a Crystalized Intelligence Composite (the picture vocabulary task and reading test), and a Fluid Intelligence Composite (the other five tasks) were also derived from these tasks.

CBCL: The Child Behavior Checklist (CBCL)[12] is a 113-item questionnaire on which caregivers rated items representing specific problems in the past six months. The CBCL was completed annually. Subscales include aggressive behavior, anxious/depressed, attention problems, rule-breaking behavior, somatic complaints, social problems, stress problems, thought problems, and withdrawn/depressed. There is a broadband scale for internalizing problems, which sums the anxious/depressed, withdrawn-depressed, and somatic complaints scores, and a broadband scale for externalizing problems, which sums rule-breaking and aggressive behavior. The total problems score is the sum of the scores of all the problem items. The CBCL also includes a set of DSM-oriented scales for depressive problems, affective problems, anxiety problems, somatic problems, ADHD problems, oppositional defiant problems, conduct problems, obsessive-compulsive problems, and sluggish cognitive tempo. Analyses examined the raw CBCL scores and included both higher-order factors (i.e., total problems and externalizing), as well as lower-order scales (e.g., DSM-5 ADHD problems), as prior work has identified both shared and distinct associations between brain metrics and psychopathology[13].

Psychotic-like Experiences: Children completed the 21-item Prodromal Questionnaire-Brief Child Version (PQ-BC)[14] in which they responded yes/no to whether they experienced a thought/feeling/experience (e.g., do familiar surroundings sometimes seem strange, confusing, threatening, or unreal to you?) before reporting whether it was distressing (yes/no) and if so, the extent (i.e., 0= no, 1 = yes [but no distress], 2–6 = yes [1 + score on distress scale]) that it bothered them. From these data, Total (i.e., the sum of endorsed items) was used as a measure of psychotic-like experiences, following evidence that the Total score is more strongly associated with psychosis risk factors[14].

Additional covariates included in supplemental analyses included:

Child Race: Parents/caregivers selected from 26 categories. Dichotomous groups were formed for the most prevalent categories of race (i.e., White, Black, Asian, Pacific Islander, Native American) with remaining participants being assigned to Other. All variables were dummy coded as non-mutually exclusive dichotomous variables; as such, participants could be coded within more than one category.

Child Ethnicity: Parents/caregivers reported whether they consider the child to be Hispanic/Latinx.

Maternal Education: Maternal education was recoded such that 12th grade, HS grad, and GED =12 years; some college and associate's degree=14 years; Bachelor's degree=16 years; Master's degree=18 years; Professional and Doctoral degrees=20 years.

Household Income: Due to low endorsement of the first five of 10 household income levels, this variable was recoded such that the first six categories were assigned a value of one (i.e., <$50,000). The subsequent categories used were coded as two ($50,000-$74,999), three ($75,000-$99,999), four ($100,000-$199,999), and five ($200,000 or more), respectively.

Balanced split-halves

The ABCD dataset was divided into two balanced splits to facilitate machine learning training and testing without data leakage. All observations from a given imaging site were placed in the same split and all possible splits (10 sites in one split, 11 in the other = $\frac{21!}{10! * 11!}$ = 352,716) were checked for balance with regards to participant age, sex, number of visits with imaging data (one or two), pubertal status, in-scanner motion, and task performance (accuracy and reaction-time during the 2-back and 0-back conditions). The most well-balanced split was also reasonably balanced with regard to participant race (the number of white and Black identifying participants did not differ) and family income (**Supplemental Table 1**).

Task effect maps

Task effect maps were computed in the whole sample. Linear mixed effect models[15] tested the effect of condition (i.e., 2-back>0-back) on activation in each region, controlling for average head motion, scanner manufacturer, and split as fixed effects, and participant, site, and family as random-intercepts. Task effect maps were computed 1) across all visits, 2) separately for each wave, and 3) separately for each run (1 and 2) of each wave.

Elastic-net machine learning

Classifier parameters ($\alpha$ and $\lambda$) were selected via grid search across 20 values for each (i.e., 400 parameter combinations)[16]. Five-fold cross-validation, repeated 10 times, was used to evaluate the performance of each parameter combination. All observations from a given imaging site were always included in the same cross-validation split. The optimal elastic-net parameters were selected via the one-se rule[17] (i.e., selecting the least complex model that achieves an accuracy within one standard error of the best-performing model). A final model was then trained using the selected parameters.

Feature importance: Logistic elastic-net regression returns the regression estimate of each feature (i.e., brain region) as a measure of that feature's contribution to the model's prediction. However, these regression estimates represent the contribution of each feature in the context of the other variables included in the model (i.e., similar to the well-known Table 2 fallacy[18]) and as such do not necessarily directly capture the underlying activation pattern that the model reflects[19]. Haufe et al.[20] describe a transformation by which the signal component that a linear decoder model has identified can be reconstructed. In the case of a decoder model that yields a single prediction (as is the case here), the Haufe Transformation reduces to simply the covariance between the original training data and the model's prediction (in the training data)[21]. Prior work has shown that the Haufe Transformation yields estimates of feature importance that are far more replicable than the original machine learning regression estimates[21].

Additional machine learning

Additional machine learning algorithms were used to compute neural signatures, and the results of these algorithms were compared to the logistic elastic-net regression used in primary analyses. As in the primary analyses, models were trained in one split, and tested in the other. Model hyperparameters were determined using a grid search with five-fold cross validation, repeated 10 times. Algorithms included three linear and three non-linear algorithms. Linear algorithms were: LASSO-PCR[22] (logistic lasso regression with a principal components preprocessing step), PLS-LDA[16,23] (partial least squares data reduction followed by linear discriminant analysis classification), and SVM[24] (support vector machine classification) with a linear kernel. Non-linear algorithms were: SVM[25] (support vector machine classification) with a radial basis function (RBF) kernel, and two ensemble tree-based methods, gradient boosted machines[26] (GBM), and Random Forest[27] (RF). Model grid search considered all combinations of the following hyperparameters: LASSO-PCR: lambda=81 values ($2x10^{-4}$ - 0.2); PLS-LDA: number of PLS component=1 - 30; SVM-linear: Cost=[.001,.25,1,5,10]; SVM-RBF: cost=[.001,.25,1,10,100], sigma=[.0001,.001,.01,1,10]; GBM: number of trees=[50,100,250,500], interaction depth=[1,3,6,10], shrinkage=[0.001,.01,0.1,1], minimum number of observations in terminal nodes=[1,10]; RF: number of variables to use=[7,13,56], number of trees=[100,250,500,1000)]. Models were then compared on three outcomes: 1) the area under the ROC curve, 2) haufe-transformed weights, and 3) predicted condition probability for the 0-back and 2-back conditions, as well as the difference between 2-back and 0-back (**Supplemental Figure 3**).

<u>Permutation tests for brain map correlations</u>

Whole-brain maps (i.e., activation, feature importance, or associations with cognition/psychopathology) were compared using a pairwise correlation. However, the significance of this correlation cannot be determined following the usual approach, as individual brain regions are non-independent - also referred to as spatial auto-correlation[28]. Here we take advantage of the fact that for all of our brain map comparisons, one or both of the maps are from tests or models in which the two task conditions, 0-back and 2-back, are compared. Thus, while the hierarchical structure of the data is complex (i.e., random effects of subject, family, and site), it is simple to generate valid permutations of the data by permuting condition labels within-subject[29]. For example, for the data [subj1={0,2}, subj2={0,2}], [{2,0},{0,2}],[{0,2},{2,0}], [{2,0},{2,0}] are all possible permutations (in addition to the original data ordering, which is also a valid result of the permutation procedure). The resulting permuted data is then used to generate a new brain map, either through linear mixed effect models or through logistic elastic net regression. In either case, the full procedure for deriving a brain map is followed, including five-fold cross-validation, repeated 10 times, across 400 parameter combinations, in the case of logistic elastic net regression. The result is a valid null brain map that exactly preserves the spatial auto-correlation of the original brain map. Repeating this procedure 10,000 times and computing the correlation between the null brain map and a true brain map each time, we obtain empirical null distributions for each correlation of interest. Significance is then computed by fitting a beta distribution to the empirical null distribution[30] (values are squared first), and using the resulting fit to compute the probability of observing an effect as large or larger than a given correlation[31]. This approach allows for the resolution of p-values smaller than 1/10000, or 0.0001. Here we refer to this approach as resulting in 'robust' p-values ($p_{robust}$), as they are robust to the effects of spatial auto-correlation.

<u>Model comparison with AIC</u>

To compare the individual difference associations of the neural signature and the linear elastic net model regression, mixed effect models were fit with maximum likelihood. The change in AIC (Akaike information criterion) between each model and the shared covariate-only model was calculated, and the difference in AIC-improvement was examined. The significance of the difference in AIC-improvement was found by performing 10,000 parametric bootstraps for each individual difference outcome.

Neural signature simulations with synthetic data

In order to further investigate what parameters influence the performance of a signature model, and how the signature approach compares to a standard linear model, analyses with synthetic simulated data were conducted. Synthetic data were generated by simulating data from a within-subjects experimental manipulation with two or more outcome variables, all of which are independent and multivariate normal. To best reflect the standard approach to analyzing task fMRI data, difference scores were then computed for each outcome variable. The individual difference measure of interest was then simulated as a normally distributed variable that is equally correlated with each of the difference scores. A signature model was trained using a logistic regression to distinguish between the two different conditions, and the difference between the prediction for the two conditions was taken. A linear model was trained using the difference scores to predict the outcome measure. The performance of these two models was then tested in an independent testing sample, the size of which was held constant (N=5,000). Parameters that were varied included: Training sample size (N= 20, 40, 80, 160, 320, 640, 1280, 2560), Number of outcome variables (n=2, 4, 6, 8, 10), Main effect of the experimental manipulation (Cohen's D, equal across all outcome, sampled uniformly from [0,1]), Correlation between the two experimental conditions (r, sampled uniformly from [0,0.9]), and total variance explained by the difference score measures in the population ($R^2$, sampled uniformly from [0.005,0.6]). A total of 72 million simulations were performed, 1 million simulations for each of the 72 unique combinations of training sample size and number of outcome variables.

To further explore the impact of the correlation between the main effect of task and regional associations on signature performance, additional simulations with more variables and more complex correlational structures were conducted. Owing to their size, a smaller number of simulations were performed. Data (n=300 variables, n=150 difference score variables) were simulated by first generating a covariance matrix with a complex structure. The eigenvalues of the N-back task data were examined, and eigenvalues were drawn from a similarly skewed distribution (gamma distribution, shape=0.05, rate=1.5). Eigenvectors were drawn from a normal distribution (mean=0, sd=1). A covariance matrix was then generated from these, and multivariate normal data were simulated using this covariance matrix. Difference scores were then created by using hierarchical clustering to identify pairs of variables with similar patterns of correlations. Correlations between the individual difference measure and the principal components of the difference scores were drawn from a normal distribution, and re-scaled to control the total population $R^2$ ($R^{2=}$0.01, 0.1) The individual difference measure of interest was then simulated as a normally distributed variable. Difference score means were then simulated as a normally distributed variable that is correlated with the vector of correlations between the individual difference measure and the difference scores themselves. This correlation (the map correlation, or map alignment) was varied (r=0, 0.2, 0.4, 0.6, 0.8, 0.9, 0.99) within each simulation round. Difference scores were then re-meaned according to the new vector of means. A signature was then created in the training data (N= 20, 40, 80, 160, 320, 640, 1280, 2560) using partial least squares (PLS) regression followed by linear discriminant analysis (LDA) (5 CV-folds, 5 repetitions) and a standard ML prediction model was trained using PLS regression (5 CV-folds, 5 repetitions). PLS was used as it is fast to train and performs relatively well. Model predictions were examined in a testing sample of N=5,000. 10,000 simulations were repeated for each sample size and population $R^2$ combination; all map correlations were examined within each simulation.

HCP MRI data acquisition and processing

Structural MRI: High-resolution (0.7-mm isotropic voxels) anatomical images were acquired using a customized Siemens Skyra 3-T scanner with a 32-channel head coil[32]. T1-weighted high resolution structural images acquired using a 3D MPRAGE sequence (FOV = 224 mm, matrix = 320, 256 sagittal slices,

TR = 2400 ms, TE = 2.14 ms, TI = 1000 ms, FA = 8°) were used in the HCP minimal pre-processing pipelines to register functional MRI data to a standard brain space.

Functional MRI: Participants completed two runs of an eight-block N-back working memory task[33]. Participants were presented with stimuli (i.e., images of faces, places, tools, or body parts) and asked to indicate whether or not they match a target image. In the low working-memory load ‘0-back’ condition, a target stimulus is presented at the beginning of each block, and participants are asked to indicate whether each subsequent stimulus matches the target image. In the high working-memory load ‘2-back’ condition, participants indicate whether a given stimulus matches the stimulus presented two screens prior. Each run consisted of four 0-back and four 2-back blocks. Behavioral measures include accuracy and reaction time in the 0-back and 2-back conditions.

2.0 mm isotropic BOLD (blood-oxygen level dependent) functional magnetic resonance images (fMRI) were acquired. Scans used multiband EPI with a slice acceleration factor of 8 (TR=720ms, TE=33.1ms, Flip Angle=52°). Preprocessing included corrections for head motion, $B_0$ distortion, gradient nonlinearities, and between-scan motion[32]. fMRI images were then registered to T1w images, initial frames were removed, and voxel time series were normalized and sampled onto the cortical surface. Time courses were temporally filtered using Gaussian-weighted linear highpass filter with a (soft) cutoff of 200 s. Effects of task on activation were computed using a general linear model. Hemodynamic response functions were modeled as square waves convolved with a two parameter gamma basis function[2]. Average activation was calculated for cortical surface-based ROIs using the Destrieux cortical atlas[9].

HCP Measures

Cognition: As in the ABCD, participants in the HCP completed the NIH toolbox, including a picture vocabulary task (language and verbal intellect), a reading test (language), a pattern matching task (processing speed), a list sorting task (working memory), a memory test (episodic memory), a response inhibition task (attention and executive function), and a card sort task (executive function and cognitive flexibility)[11]. Participants additionally completed measures of sustained attention (Short Penn Continuous Performance Test), verbal episodic memory (Penn Word Memory Test), spatial orientation (Variable Short Penn Line Orientation Test), and fluid intelligence (Penn Progressive Matrices)[34,35].

**Supplemental Results**

Comparing neural signature associations to individual regional associations

Signature associations were significantly larger than all regional associations for all of the task performance and cognitive measures examined ($p<0.05$ bonferroni-corrected, **Supplemental Methods & Data**). Signature associations were additionally significantly larger ($p<0.05$ bonferroni-corrected) than all or most regional associations with many psychopathology measures, including total problems, psychotic-like experiences, and externalizing-related measures including attention problems, rule-breaking behavior, and conduct problems (92-100% of regions; **Supplemental Data**). Signature associations were significantly larger for only some or none of the remaining outcomes, including depression and anxiety problems (0-43% of regions; **Supplemental Data**).

Faces vs Places Classifier

To test whether the performance of the neural signature may be attributable to unadjusted confounding or sensitivity to overall task engagement, a second signature was constructed, decoding activation to the face and place stimuli used in the task (**Supplemental Methods**). This faces signature (i.e., faces>places) had high decoding accuracy and was reliable (**Supplemental Figures 13-14**), but there were minimal significant associations with behavior/psychopathology (**Supplemental Figure 15**). As the regional importance map suggests this signature is driven by differential processing in visual processing regions (**Supplemental Figure 13**), it is not surprising that associations with cognition or mental health would be minimal[36–38]. However, this null result does suggest that associations with the emotional n-back signature do not purely reflect uncontrolled confounding (e.g., head motion causing scans to be less accurately classified) or overall task engagement.

| Variable | Split1 (n=4,215) | Split2 (n=4,836) | $X^2$/t | p |
|---|---|---|---|---|
| Number of visits (two) | 2060 (48.87) | 2420 (50.04) | 1.18 | 0.28 |
| Number of family members* | 2.14 (1) | 2.17 (0.98) | 0 | 1 |
| Age (weeks)* | 130.28 (14.18) | 130.5 (14.39) | 0.87 | 0.38 |
| Sex (Female) | 2015 (47.81) | 2357 (48.74) | 0.75 | 0.39 |
| Pubertal status* | 1.78 (0.91) | 1.78 (0.89) | -0.51 | 0.61 |
| Maternal Education (years)* | 15.5 (2.36) | 15.59 (2.43) | 0 | 1 |
| **Household income, $** | | | | |
| ≤ 49,999 | 1148 (27.24) | 1278 (26.43) | 0.71 | 0.4 |
| 50,000 – 74,999 | 622 (14.76) | 636 (13.15) | 4.72 | **0.03** |
| 75,000 – 99,999 | 659 (15.63) | 714 (14.76) | 1.26 | 0.26 |
| 100,000 – 199,999 | 1301 (30.87) | 1588 (32.84) | 3.94 | **0.047** |
| ≥ 200,000 | 485 (11.51) | 620 (12.82) | 3.51 | 0.061 |
| **Race/ethnicity** | | | | |
| White | 3361 (79.74) | 3810 (78.78) | 1.19 | 0.28 |
| Black | 722 (17.13) | 826 (17.08) | 0 | 0.97 |
| Hispanic | 807 (19.15) | 904 (18.69) | 0.27 | 0.6 |
| Native American | 70 (1.66) | 236 (4.88) | 70.47 | **4.67E-17** |
| Pacific Islander | 33 (0.78) | 23 (0.48) | 2.98 | 0.08 |
| Asian | 235 (5.58) | 351 (7.26) | 10.26 | **1.36E-03** |
| Other | 208 (4.93) | 336 (6.95) | 15.8 | **7.03E-05** |
| Average head motion (mm)* | 0.29 (0.32) | 0.28 (0.31) | -1.12 | 0.26 |
| 2-back Accuracy (%)* | 0.83 (0.09) | 0.82 (0.09) | -1.15 | 0.25 |
| 2-back RT (ms)* | 1009.59 (132.7) | 1010.96 (129.94) | 0.49 | 0.62 |
| 0–back Accuracy (%)* | 0.89 (0.09) | 0.89 (0.09) | -0.29 | 0.77 |
| 0-back RT (ms)* | 862.33 (127.26) | 862.77 (127.96) | 0.19 | 0.85 |

**Supplemental Table 1. Comparison between splits**. Split 1 included 10 imaging sites and 6,248 observations from 4,215 participants. Split 2 included 11 imaging sites and 7,729 observations from 4,836 participants. Continuous variables (*) were compared with an t-test, values reflect the mean and SD (standard deviation) for each group. Binary variables were compared with a chi-squared test ($X^2$), values reflect the N (sample size) and group percentage. RT=Reaction Time. All tests were two-sided. Bold=p<0.05, uncorrected for multiple comparisons.

| Condition | Type | reliability | lower-CI | upper-CI | p |
|---|---|---|---|---|---|
| 0-back | baseline - split-half | 0.293 | 0.270 | 0.315 | $4.81 \times 10^{-131}$ |
| 0-back | follow up - split-half | 0.283 | 0.258 | 0.307 | $1.37 \times 10^{-27}$ |
| 0-back | longitudinal stability | 0.167 | 0.138 | 0.196 | $1.37 \times 10^{-27}$ |
| 2-back | baseline - split-half | 0.514 | 0.496 | 0.532 | $<1.0 \times 10^{-320}$ |
| 2-back | follow up - split-half | 0.511 | 0.491 | 0.531 | $2.28 \times 10^{-93}$ |
| 2-back | longitudinal stability | 0.310 | 0.282 | 0.338 | $2.28 \times 10^{-93}$ |
| 2-back>0-back | baseline - split-half | 0.474 | 0.454 | 0.493 | $<1.0 \times 10^{-320}$ |
| 2-back>0-back | follow up - split-half | 0.431 | 0.408 | 0.454 | $1.95 \times 10^{-81}$ |
| 2-back>0-back | longitudinal stability | 0.287 | 0.259 | 0.315 | $1.95 \times 10^{-81}$ |

**Supplemental Table 2. Neural signature reliability.** Baseline=baseline wave, follow up=follow up wave 2. CI=Confidence Interval. CIs are the 95% confidence interval. P-values smaller than the range of values that can be displayed in R are reported as $p<1.0 \times 10^{-320}$.

| Variable | Estimate | SE | t | p | lower-CI | upper-CI |
|---|---|---|---|---|---|---|
| 2-Back Accuracy | 0.338 | 0.007 | 45.655 | $<1.0x10^{-320}$ | 0.324 | 0.353 |
| 0-Back Accuracy | 0.459 | 0.008 | 60.665 | $<1.0x10^{-320}$ | 0.444 | 0.474 |
| Total Composite | 0.310 | 0.011 | 27.974 | $2.2x10^{-163}$ | 0.289 | 0.333 |
| Fluid Intelligence | 0.254 | 0.012 | 21.955 | $1.8x10^{-103}$ | 0.231 | 0.277 |
| Picture Vocabulary | 0.174 | 0.008 | 21.354 | $2.3x10^{-99}$ | 0.158 | 0.191 |
| List Sorting Working Memory | 0.242 | 0.012 | 20.881 | $4.29x10^{-94}$ | 0.220 | 0.266 |
| Crystalized Intelligence | 0.162 | 0.008 | 20.095 | $4.39x10^{-88}$ | 0.146 | 0.178 |
| Picture Sequence Memory | 0.153 | 0.009 | 17.023 | $2.74x10^{-64}$ | 0.136 | 0.171 |
| Reading Recognition | 0.139 | 0.008 | 16.960 | $9.18x10^{-64}$ | 0.123 | 0.155 |
| Flanker Inhibitory Control | 0.133 | 0.009 | 14.275 | $7.18x10^{-46}$ | 0.115 | 0.151 |
| Dimensional Change Card Sort | 0.161 | 0.012 | 13.581 | $1.66x10^{-41}$ | 0.138 | 0.185 |
| Pattern Comparison | 0.099 | 0.009 | 11.317 | $1.5x10^{-29}$ | 0.082 | 0.117 |
| Attention Prob (CBCL) | -0.076 | 0.008 | -9.489 | $2.77x10^{-21}$ | -0.092 | -0.060 |
| Social Prob (CBCL) | -0.079 | 0.008 | -9.381 | $7.66x10^{-21}$ | -0.096 | -0.063 |
| DSM-5 ADHD Prob (CBCL) | -0.065 | 0.008 | -8.018 | $1.17x10^{-15}$ | -0.081 | -0.049 |
| DSM-5 Conduct Prob (CBCL) | -0.065 | 0.008 | -7.653 | $2.1x10^{-14}$ | -0.081 | -0.048 |
| Psychotic-like Experiences (PQ-BC) | -0.064 | 0.009 | -7.250 | $4.39x10^{-13}$ | -0.081 | -0.046 |
| Rule-breaking Behavior (CBCL) | -0.061 | 0.008 | -7.134 | $1.03x10^{-12}$ | -0.077 | -0.044 |
| Sluggish Cog Tempo (CBCL) | -0.052 | 0.009 | -5.963 | $2.53x10^{-9}$ | -0.069 | -0.035 |
| Stress Prob (CBCL) | -0.049 | 0.008 | -5.909 | $3.54x10^{-9}$ | -0.066 | -0.033 |
| Total Problems (CBCL) | -0.045 | 0.008 | -5.801 | $6.75x10^{-9}$ | -0.060 | -0.030 |
| Externalizing Factor (CBCL) | -0.042 | 0.008 | -5.283 | $1.3x10^{-7}$ | -0.058 | -0.027 |
| DSM-5 Depression Prob (CBCL) | -0.039 | 0.009 | -4.567 | $4.98x10^{-6}$ | -0.056 | -0.023 |
| Aggressive Behavior (CBCL) | -0.036 | 0.008 | -4.410 | $1.04x10^{-5}$ | -0.052 | -0.020 |
| DSM-5 Opposit Defiant Prob (CBCL) | -0.035 | 0.008 | -4.258 | $2.08x10^{-5}$ | -0.052 | -0.019 |
| DSM-5 Anxiety Prob (CBCL) | -0.033 | 0.008 | -3.892 | $9.98x10^{-5}$ | -0.050 | -0.016 |
| DSM-5 Somatic Prob (CBCL) | -0.030 | 0.009 | -3.395 | $6.88x10^{-4}$ | -0.047 | -0.013 |
| Somatic Prob (CBCL) | -0.030 | 0.009 | -3.387 | $7.09x10^{-4}$ | -0.047 | -0.012 |
| Internalizing Factor (CBCL) | -0.026 | 0.008 | -3.179 | 0.001 | -0.043 | -0.010 |
| Obsess-comp Prob (CBCL) | -0.026 | 0.009 | -3.069 | 0.002 | -0.043 | -0.009 |
| Thought Prob (CBCL) | -0.025 | 0.009 | -2.892 | 0.004 | -0.041 | -0.008 |
| Anxious/Depressed (CBCL) | -0.024 | 0.008 | -2.887 | 0.004 | -0.041 | -0.008 |
| Withdrawn/Depressed (CBCL) | -0.022 | 0.009 | -2.571 | 0.010 | -0.040 | -0.005 |

**Supplemental Table 3. Individual difference associations with the neural signature.** CI= Confidence Interval. CIs are the 95% confidence interval. CBCL=Child Behavior Checklist. PQ-BC=Prodromal Questionnaire-Brief Child Version. Prob=Problems.

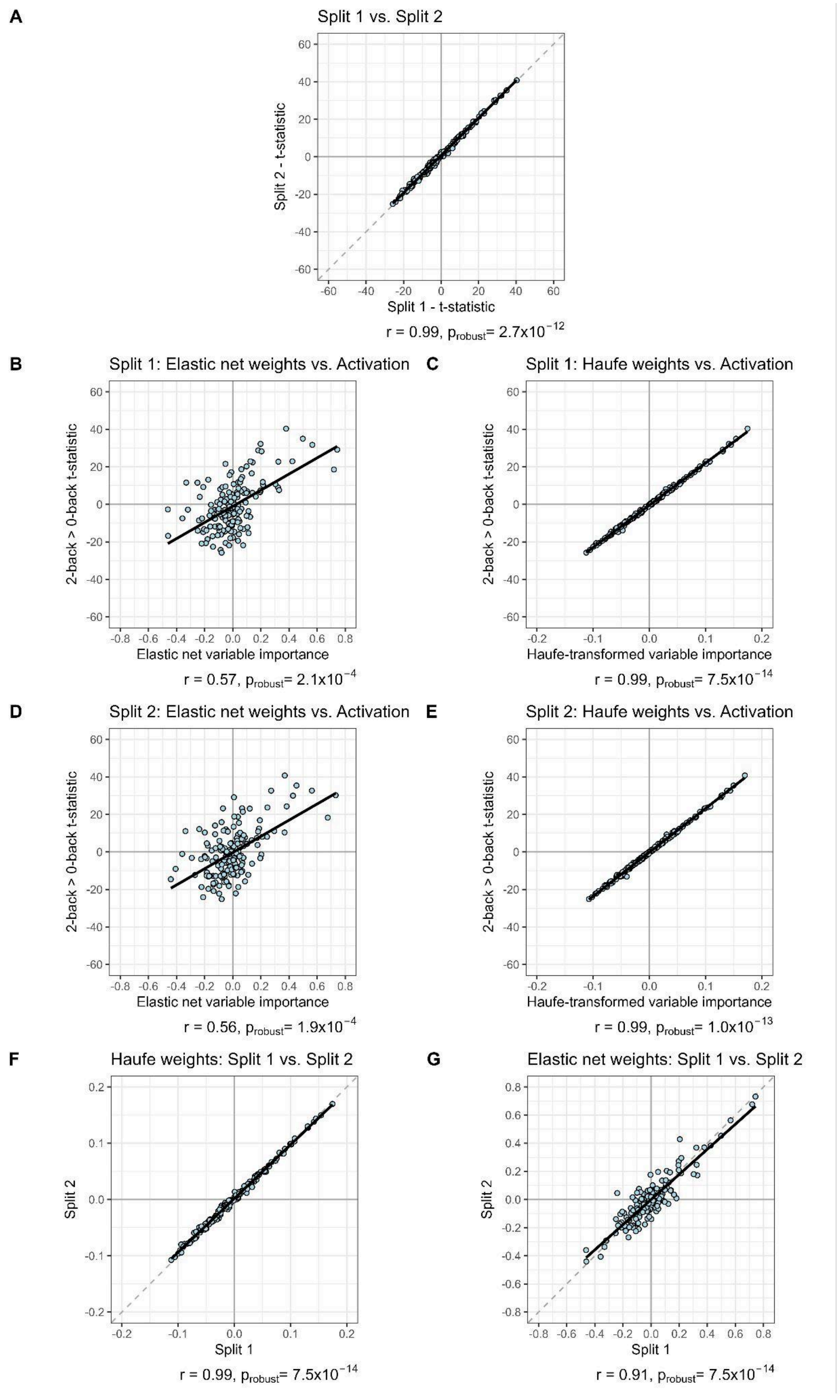


**Supplemental Figure 1**. **Comparisons across splits and measures. A)** Task similarity across splits. The main effect of the task (2-back>0-back) is highly similar across the two independent splits. **B-E)** Similarity between measures of variable importance and task activation in each of the two splits. **F-G)** Similarity in variable importance across the two splits.

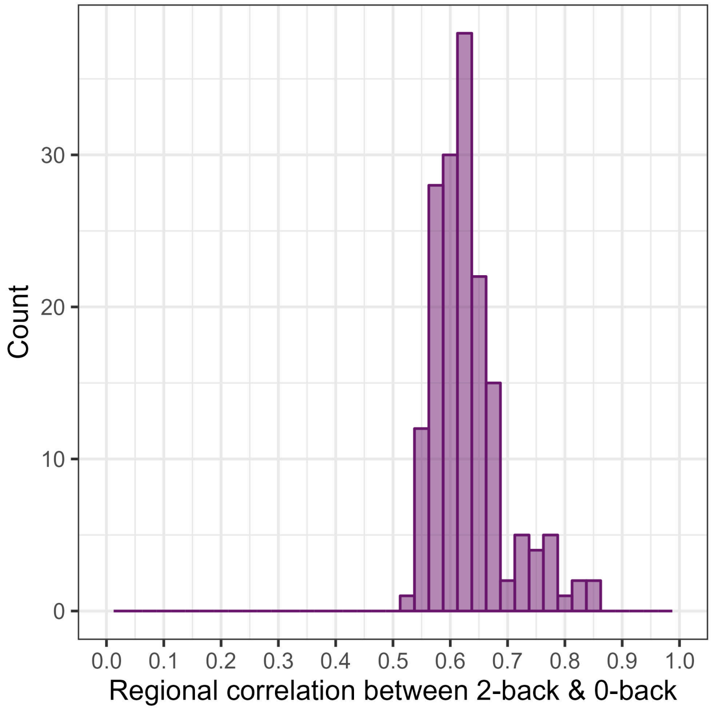


**Supplemental Figure 2**. **Regional activation is highly similar between the 0-back and 2-back conditions.** Correlation between activation in the 0-back and 2-back conditions. Each observation is a single region.

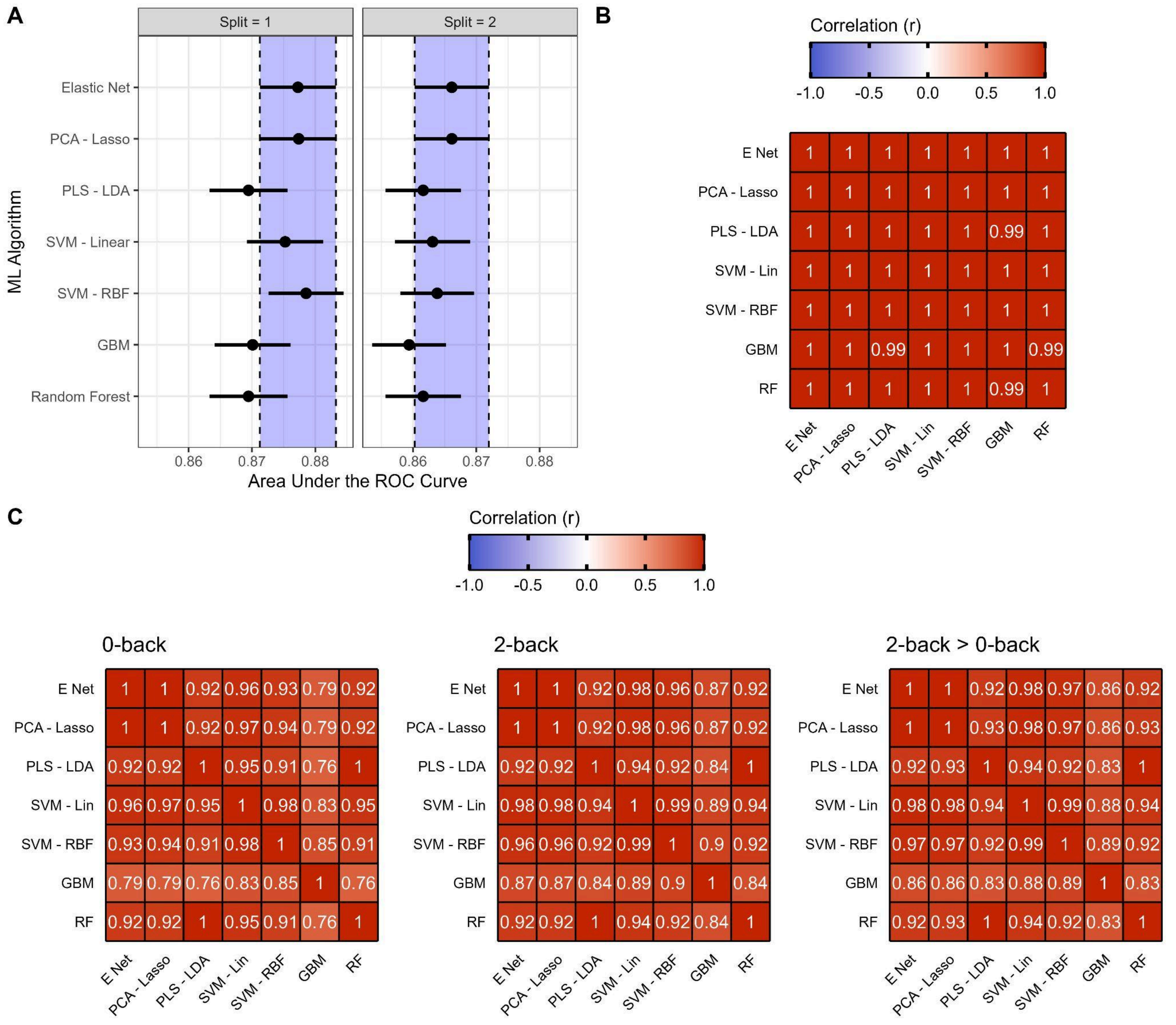


**Supplemental Figure 3**. **Comparison of machine learning algorithms. A)** Out-of sample classification performance for each of the seven algorithms. ROC=Receiver operating curve. Horizontal lines reflect the 95% confidence interval. Dashed vertical lines and the shaded region denote the 95% confidence interval of the elastic-net classifier. **B)** Haufe-transformed variable weight similarity (correlation) between the seven models. **C)** Neural signature similarity for the 0-back, 2-back, and 2-back>0-back conditions for each of the seven models. E net= Elastic net. PCA=Principal components analysis. PLS=Partial least squares. LDA=Linear discriminant analysis. SVM=Support vector machines. Lin=Linear. RBF=Radial basis function. GBM=Gradient boosted machines. RF=Random forest.

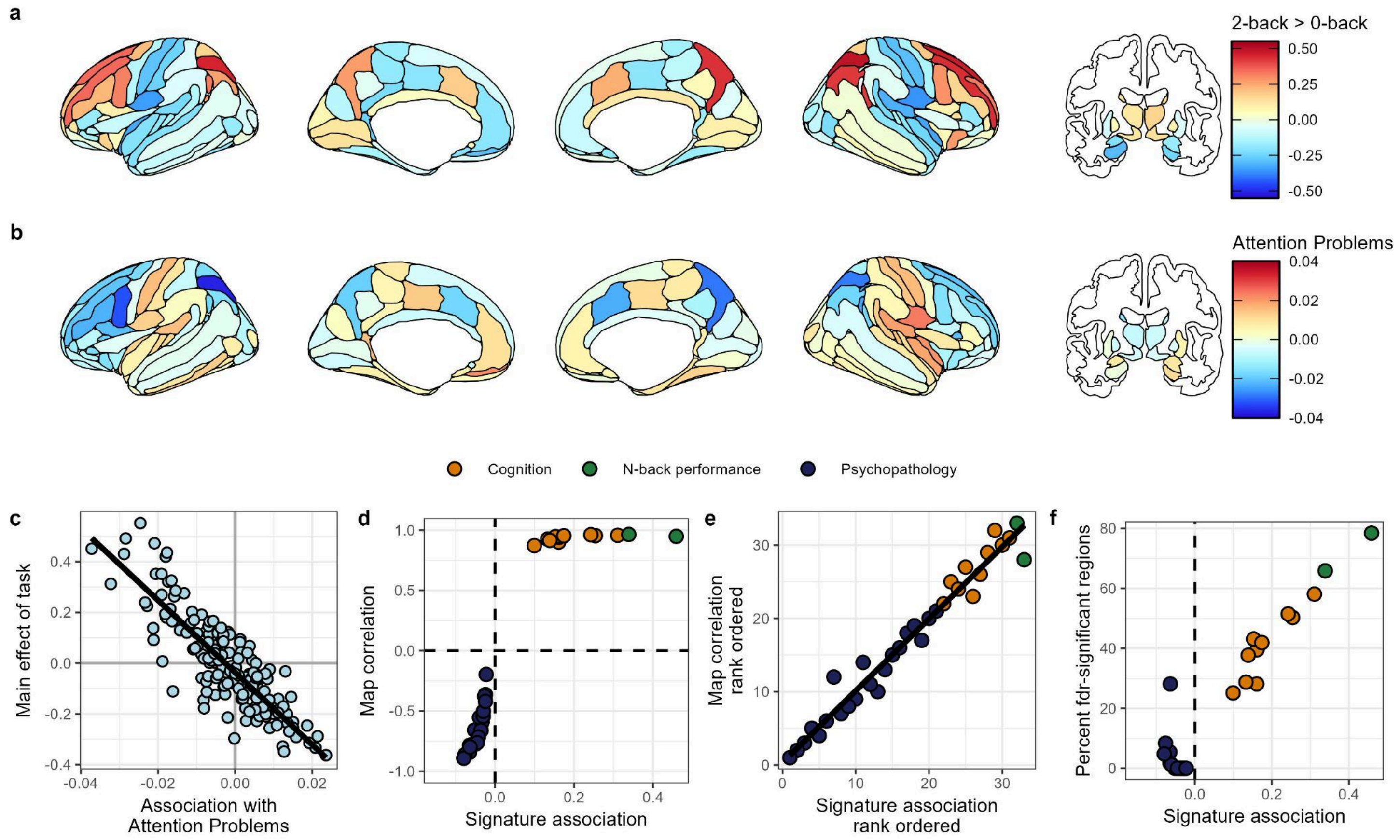


**Supplemental Figure 4**. **Neural signature associations reflect spatial alignment with the main effect of task**. **A)** Main effect of task, 2-back>0-back, colored by the standardized regression estimate. **B)** An example regional association map for attention problems (association of the 2-back>0-back regional contrast with the CBCL attention problems score), finding that attention problems are associated with less activation of key fronto-parietal working memory regions. The map is colored by the standardized regression estimate. **C)** Example spatial map correlation, between attention problems regional associations and the 2-back>0-back main effect of task, standardized regression estimates (r=-0.86, $p_{robust}$=1.7x$10^{-4}$; plots for all outcomes are shown in **Supplemental Figures 10&11**). Attention problems are associated with less activation in regions that are activated by the 2-back condition (relative to 0-back), and conversely, more activation in regions that are deactivated by the 2-back condition. **D)** Neural signatures associations (regression estimates from **Figure 3**) and C **(**the spatial map correlation) for each measure. **E)** Rank-ordered neural signature associations and rank-ordered map correlations ($\varrho$=0.98, $p_{permutation}$=1.0x$10^{-4}$). **F)** Neural signatures associations and the percent of individual regional associations that survive false discovery rate (fdr) multiple test correction. **D-F)** Colors reflect the type of individual difference measure (orange=Cognition, green=N-back performance, purple=Psychopathology).

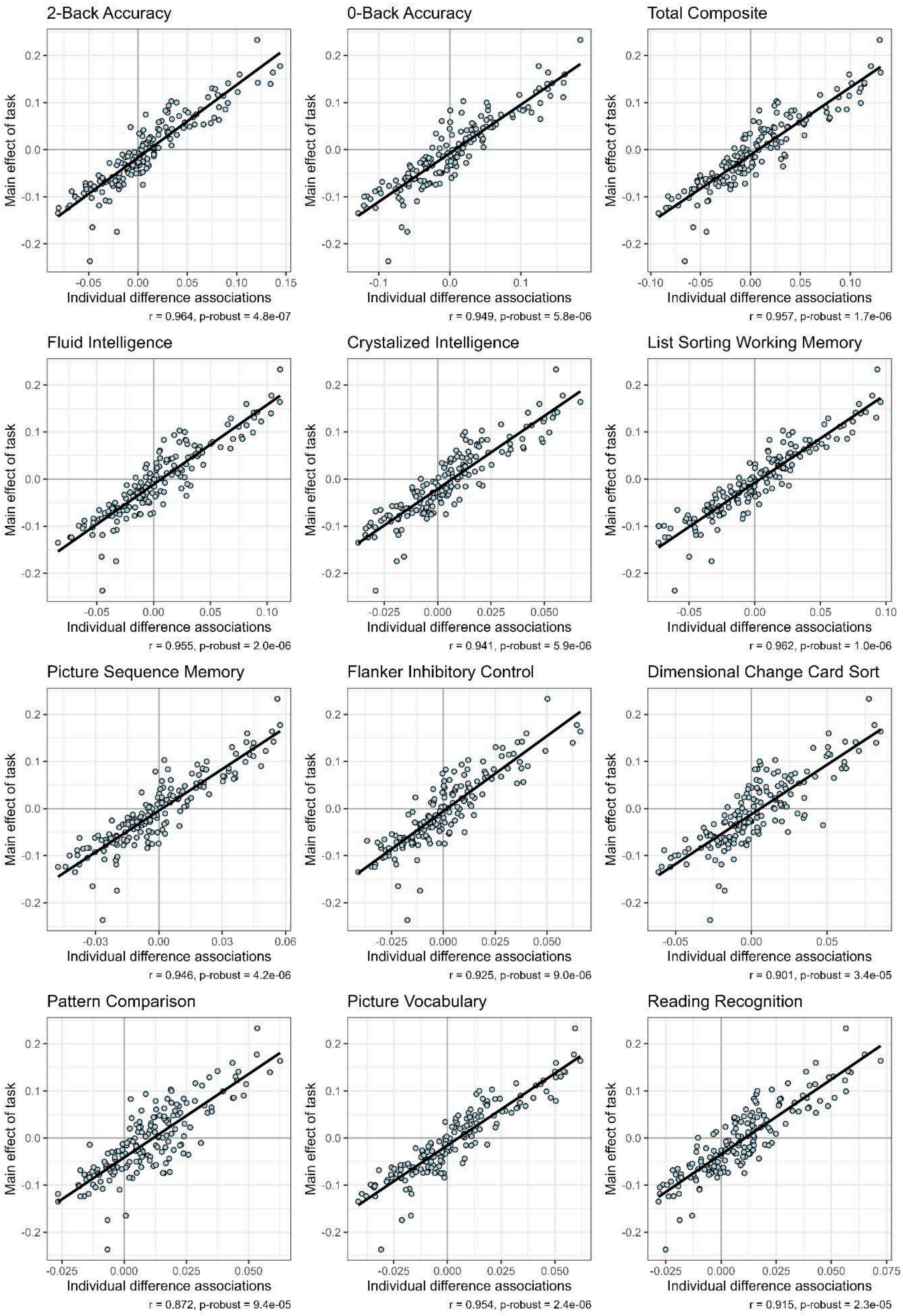


**Supplemental Figure 5**. **Spatial map correlations with task performance and cognition.** Spatial map correlations between individual difference measure associations with task activation and the 2-back>0-back main effect of task, standardized regression estimates. Pearson correlation coefficient (r) and the robust p-value are shown below each plot.

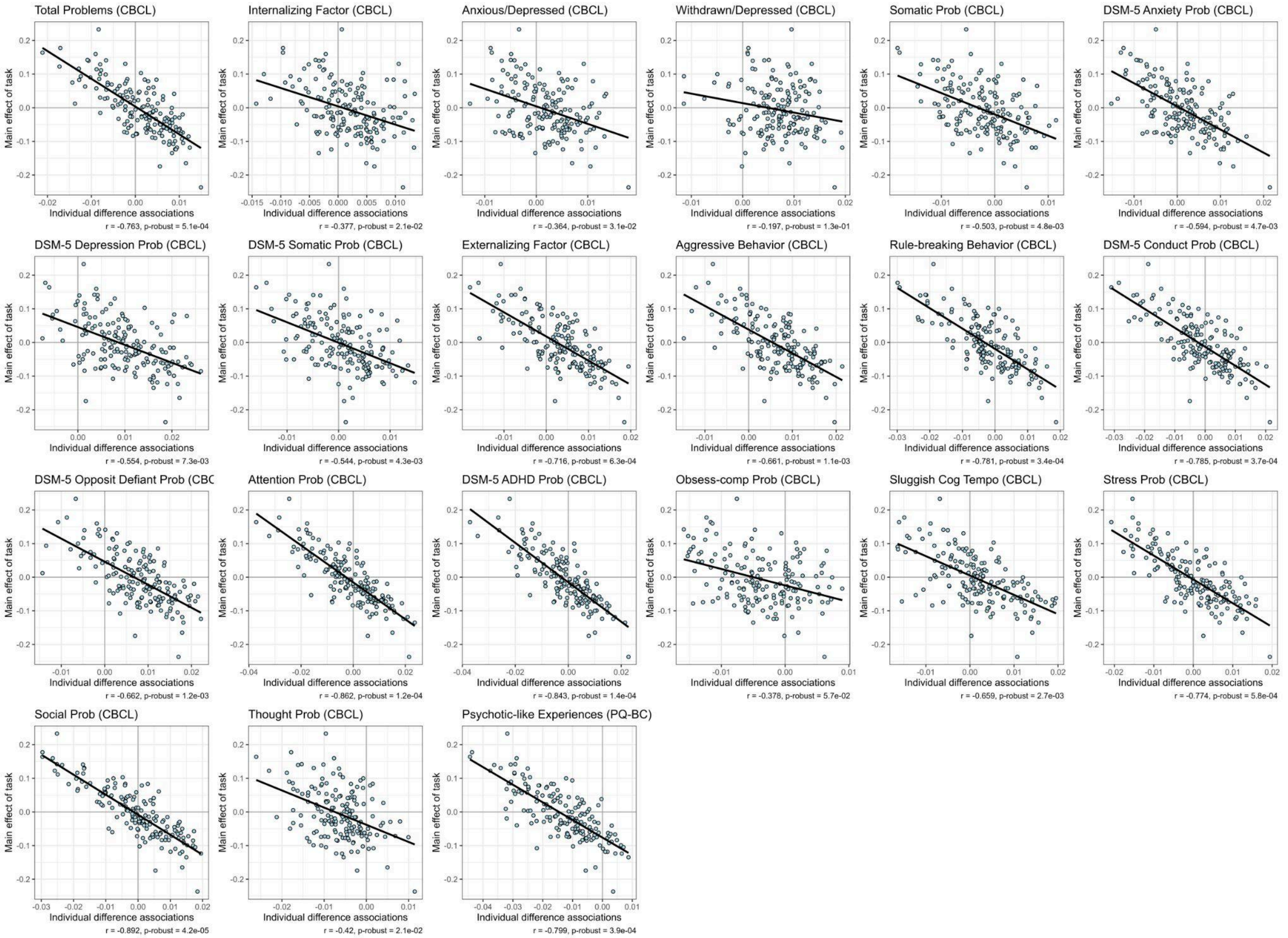


**Supplemental Figure 6**. **Spatial map correlations with psychopathology.** Spatial map correlations between individual difference measure associations with task activation and the 2-back>0-back main effect of task, standardized regression estimates. Pearson correlation coefficient (r) and the robust p-value are shown below each plot.

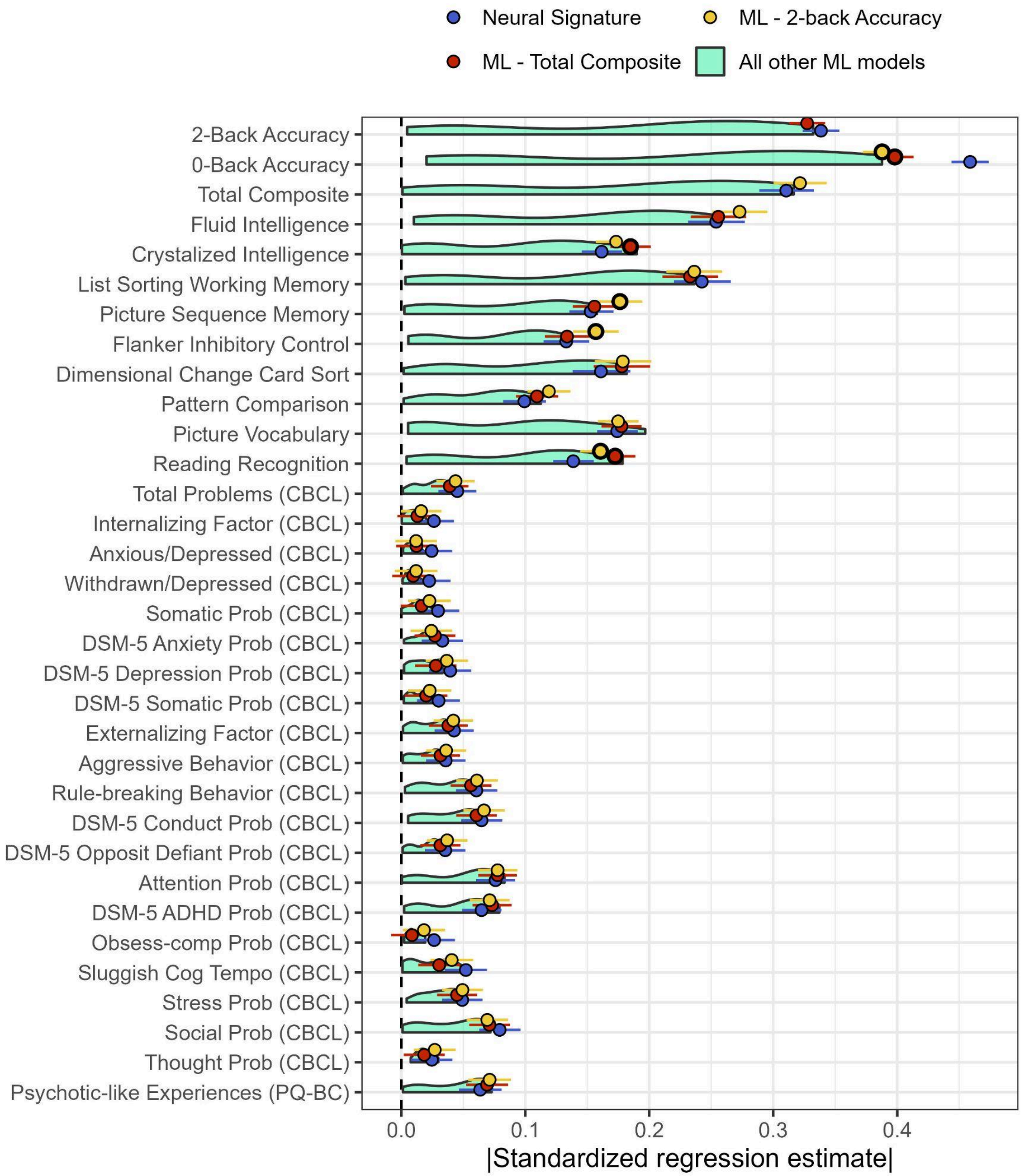


**Supplemental Figure 7**. **Generalizability of ML predictions to other outcomes**. The absolute value of the standardized regression estimate from models predicting outcomes that they were not trained on. The neural signature (blue), and ML models for 2-back accuracy (yellow) and the Total Composite score (red) are shown as points with their associated 95% confidence intervals. These two ML models are highlighted as they were the best performing across all outcomes, besides the neural signature. All other ML models are shown in light green. Bolded points indicate the model fit statistics for ML model significantly differs from the neural signature after false discovery rate correction for multiple comparisons at $p<0.05$.

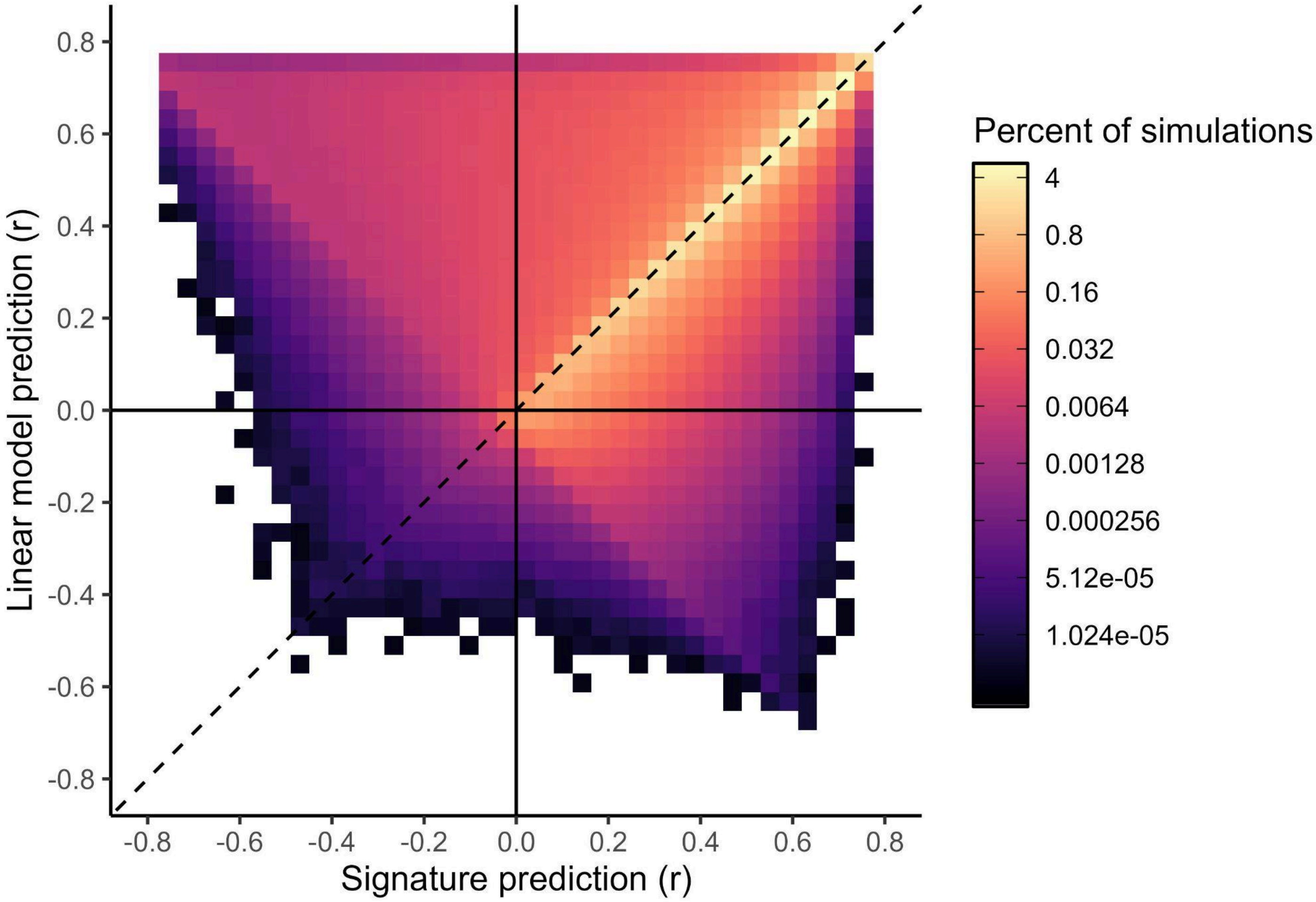


**Supplemental Figure 8**. **Comparison of signature and linear model performance across simulations**. Colors reflect the percent of the 72 million simulations that achieved a given performance level (log-scale). Equal performance is indicated by the dashed line (the diagonal). The neural signature and linear models achieved similar performance across the majority of simulations (i.e., 55% of results lie along the diagonal in the top-right quadrant).

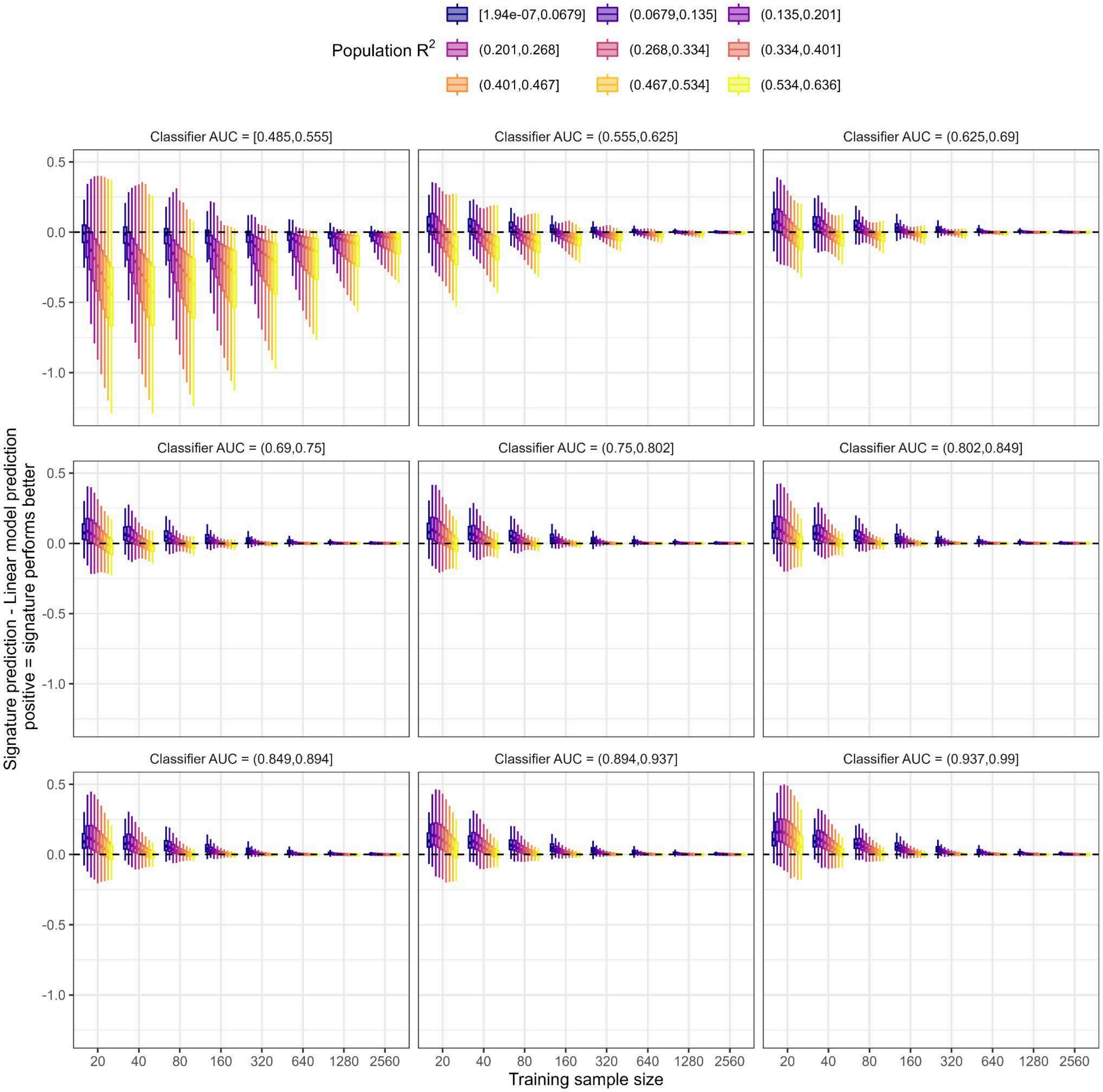


**Supplemental Figure 9**. **Variables influencing simulation model performance.** The y-axis shows the difference between neural signature and linear model performance in synthetic data. Positive values indicate better performance with the signature, negative values indicate better performance with a linear model. Data are presented as box-plots, showing the distribution of performance values simulation results. The size of the training sample is shown along the x-axis. Box-plots are colored by population $R^2$, which was approximated by the full model $R^2$ from a linear regression fit in the test sample with all variables. Each sub-panel shows results for a range of classifier performance (AUC) values, where levels were determined by splitting the distribution into nine equally-sized groups.

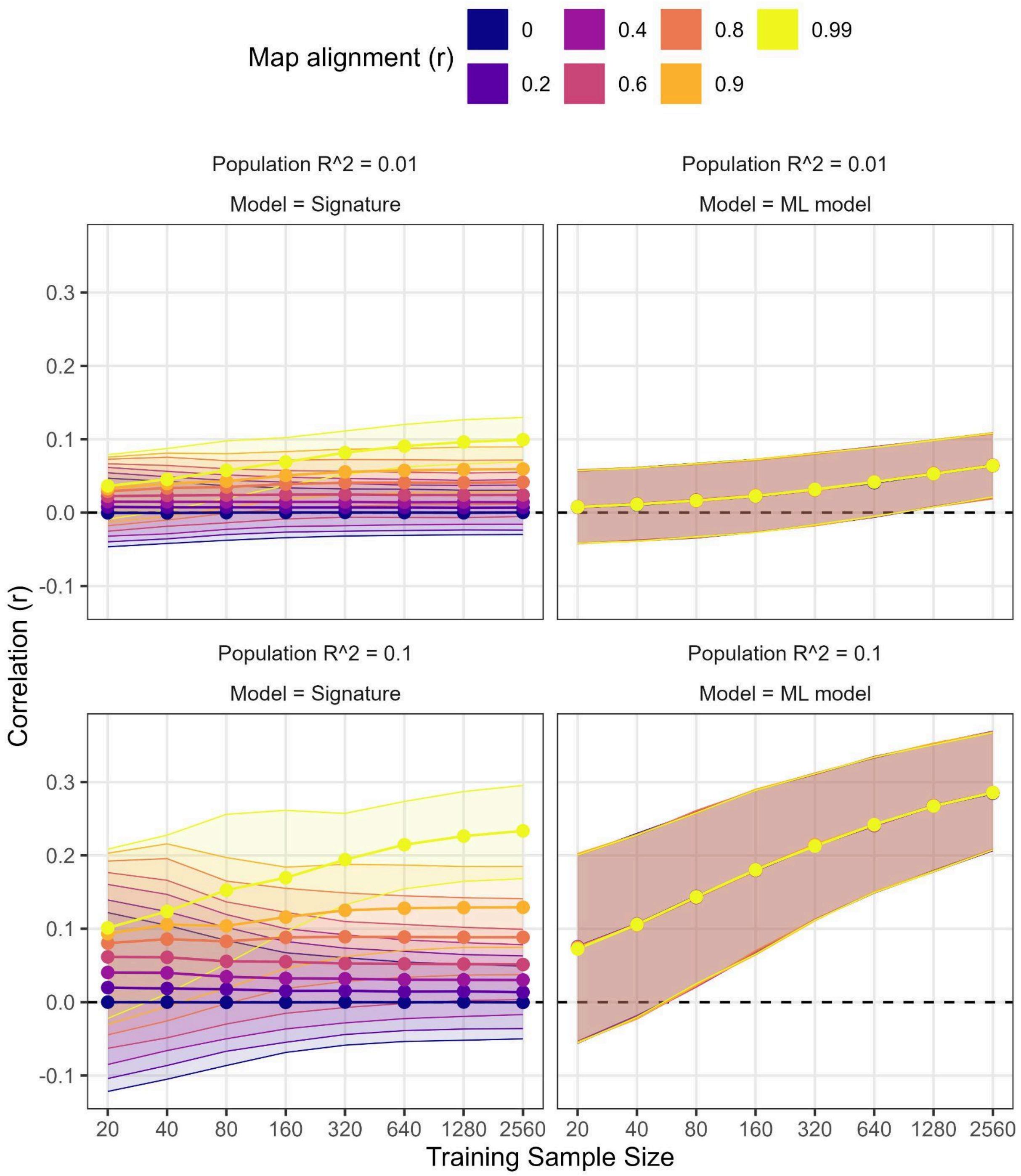


**Supplemental Figure 10**. **Influence of map alignment on simulation model performance.** Data are fully synthetic. The y-axis reflects the correlation between the model's prediction and the simulated outcome variable. Performance for the signature approach and a standard ML model (PLS regression) are shown separately. Analyses were repeated for medium and large effects, population $R^2$=0.01 & 0.1. Colors reflect the Map alignment, the correlation between the regional difference between conditions (i.e., main effect of task) and regional associations with the simulated outcome variable. This correlation was varied between r=0-0.99 across simulations. Points and the shaded region represent the median and 95% CI for the correlation at each sample size, across 10,000 simulations per sample size.

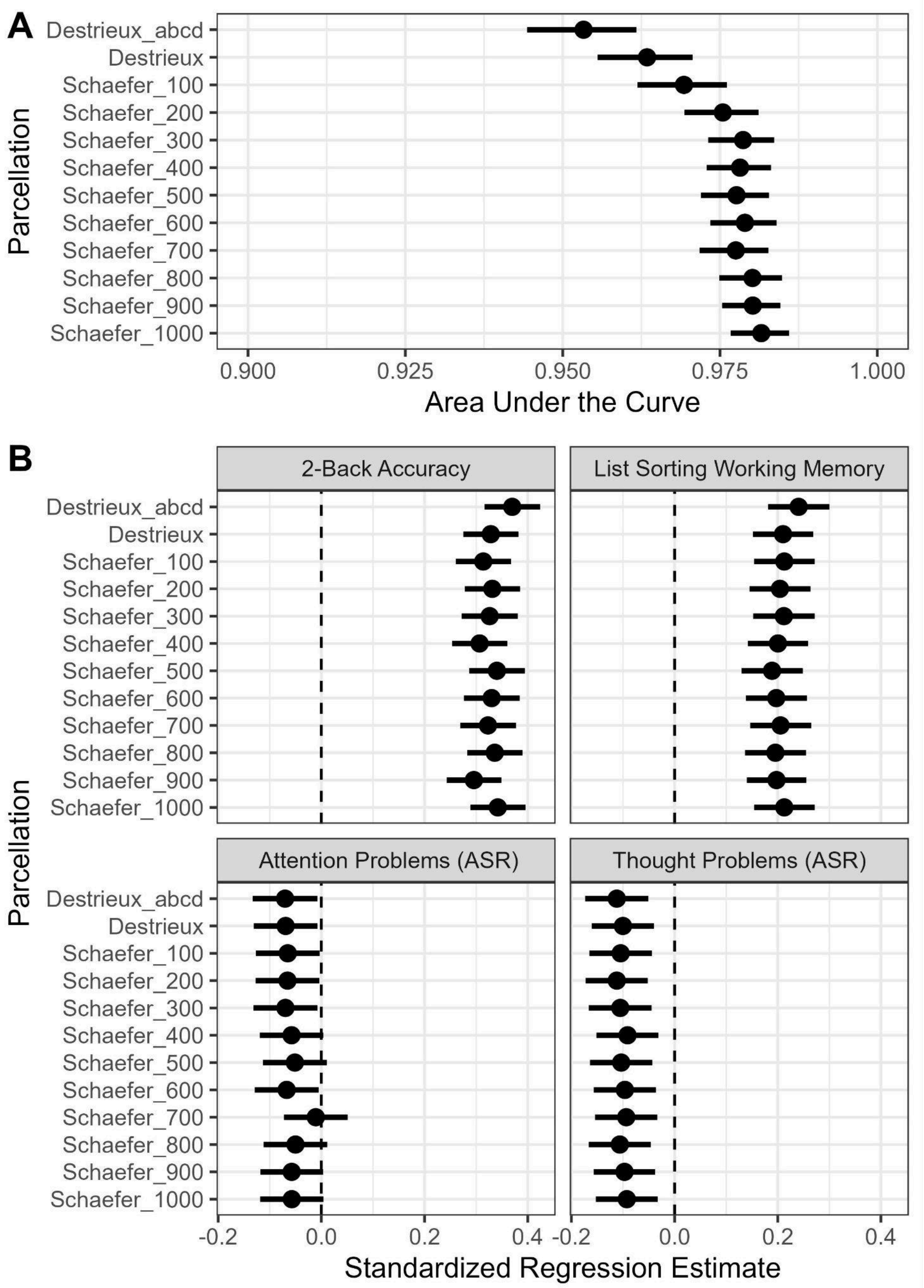


**Supplemental Figure 11**. **Influence of parcellation resolution on neural signature performance.** Using the Human Connectome Project (HCP), N-back fMRI data was parcellated using the Destrieux atlas, as well as Schaefer atlas' with 100-1,000 regions. In addition to applying the signature trained in ABCD (Destrieux_abcd), signatures were trained within HCP using each parcellation. The HCP was split into 5 balanced splits (balanced on age, sex, task performance, and in-scanner movement), keeping family members in the same split. Elastic-net decoders were then trained as in the ABCD, using 5-fold nested cross-validation, with 10 repeats per internal loop (i.e., train in 4 splits, test in the fifth split, repeat five times, once for each split). **A)** Area under the curve for each parcellation, with associated confidence interval. **B)** Individual difference associations with four key outcomes. These analyses additionally included HCP split (5 levels) as a dummy-coded variable.

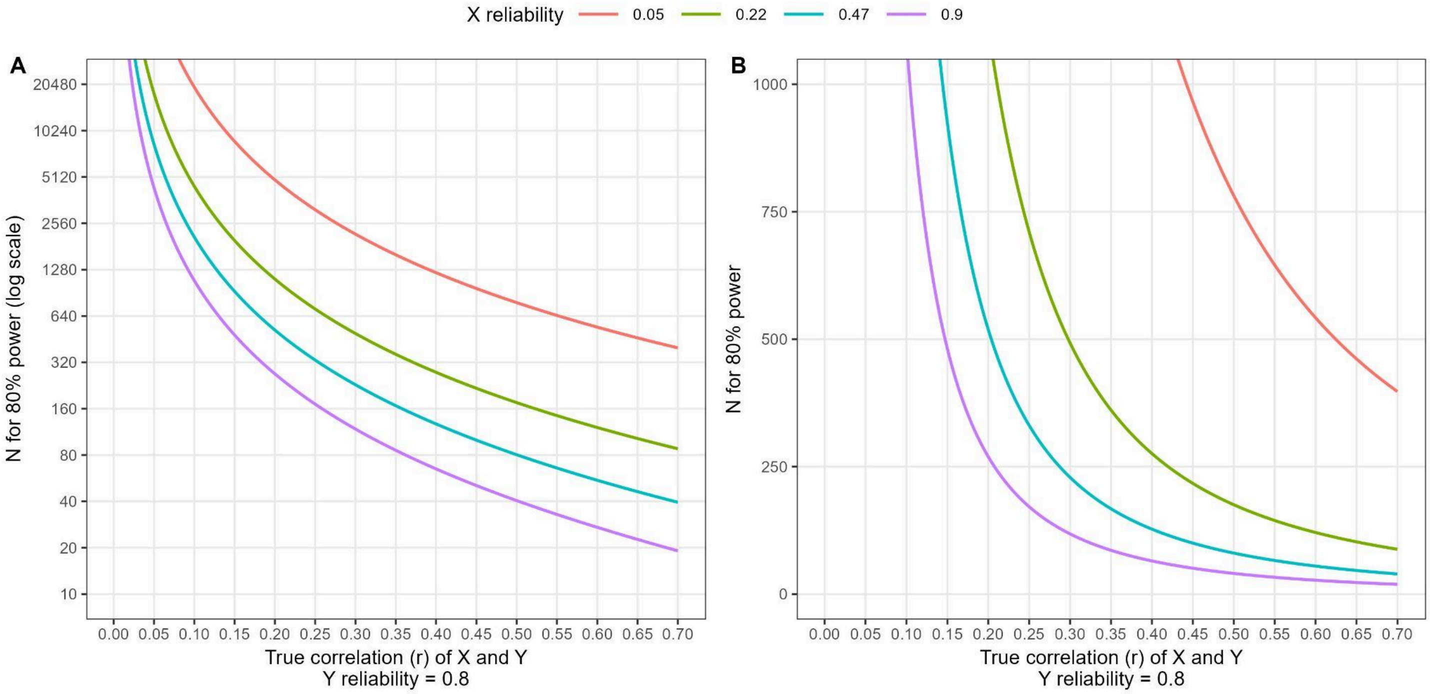


**Supplemental Figure 12**. **Greater reliability reduces the required sample size for individual difference studies.** The required sample size (y-axis; **A)** sample size is log-scale; **B**) sample size is linear scale) to detect a given true correlation (x-axis) at 80% power and alpha=0.05. Lines show the effect of X reliability at three levels: 0.05 (red) - the median regional reliability in the ABCD study; 0.22 (green) - the median regional reliability in the HCP; 0.47 (blue) - the reliability of the neural signature; and 0.9 (purple) - an ‘excellent’ reliability. The reliability of Y is held to 0.8 throughout.

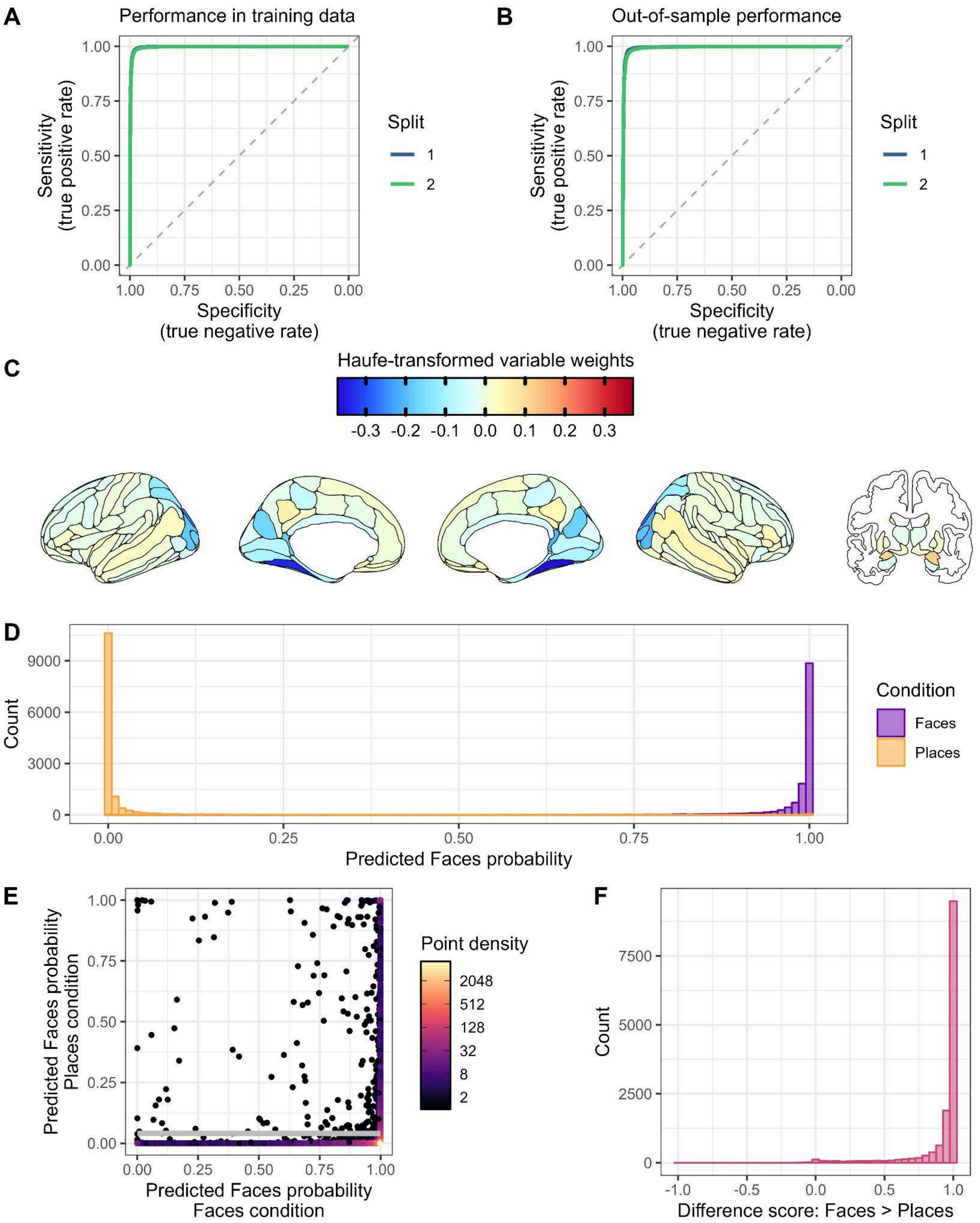


**Supplemental Figure 13**. **A neural signature of Faces vs Places. A&B**) An elastic-net classifier distinguishing face and place stimuli was trained. Model performance - area under the receiver operating characteristic curve (AUC) - was comparable across balanced split-halves in both the training splits (**A;** Split 1 AUC=0.998; Split 2 AUC=0.996) and testing splits (**B;** Split 1 AUC=0.994; Split 2 AUC= 0.993). **C)** Visualization of the haufe-transformed variable importance of each brain region, averaged across splits. Variables with larger absolute weights contribute more to the model's predictions. **D)** Histogram of model predictions for each MRI scan for each of the two conditions (purple: Places; orange; Faces), combined across data splits. Values reflect the predicted probability that a scan is from the Faces condition. **E)** Model predictions for the two conditions for each individual scan (points), colored by density (number of overlapping points). The white line reflects the correlation between predictions for the two conditions. **F)** Histogram of the difference between the model predictions for the two conditions. Values reflect how separable the two conditions are for each scan.

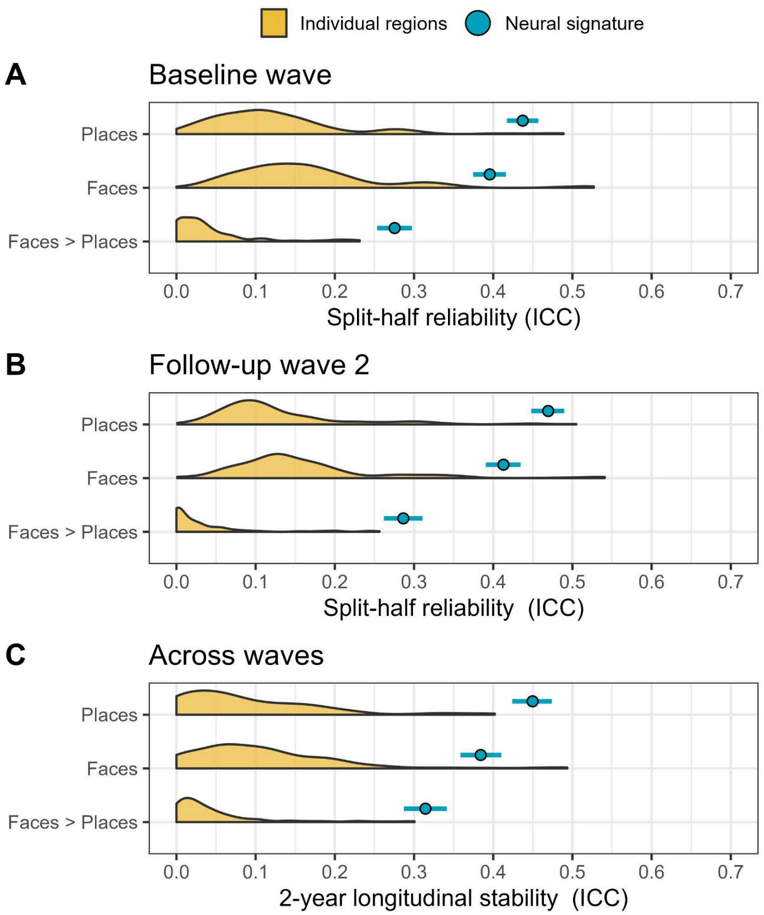


**Supplemental Figure 14. Reliability of the Faces Neural Signature and Individual Regional Activation**. The split-half reliability (comparing runs within one scan) and longitudinal stability (comparing two scans) was examined for neural signature predictions during the places condition, faces condition, and their difference (faces > places). Blue points are the neural signatures, with 95% confidence intervals indicated by horizontal lines. ICC=intraclass correlation coefficient. Shaded yellow regions represent the distribution of reliabilities across all regions (n=167) for each of the corresponding conditions and contrasts. Analyses were restricted to unrelated participants. **A)** Split-half reliability in the baseline session, N=6,551. **B)** Split-half reliability in the year 2 follow up, N=5,386. **C)** Longitudinal stability across the two waves, N=4,111.

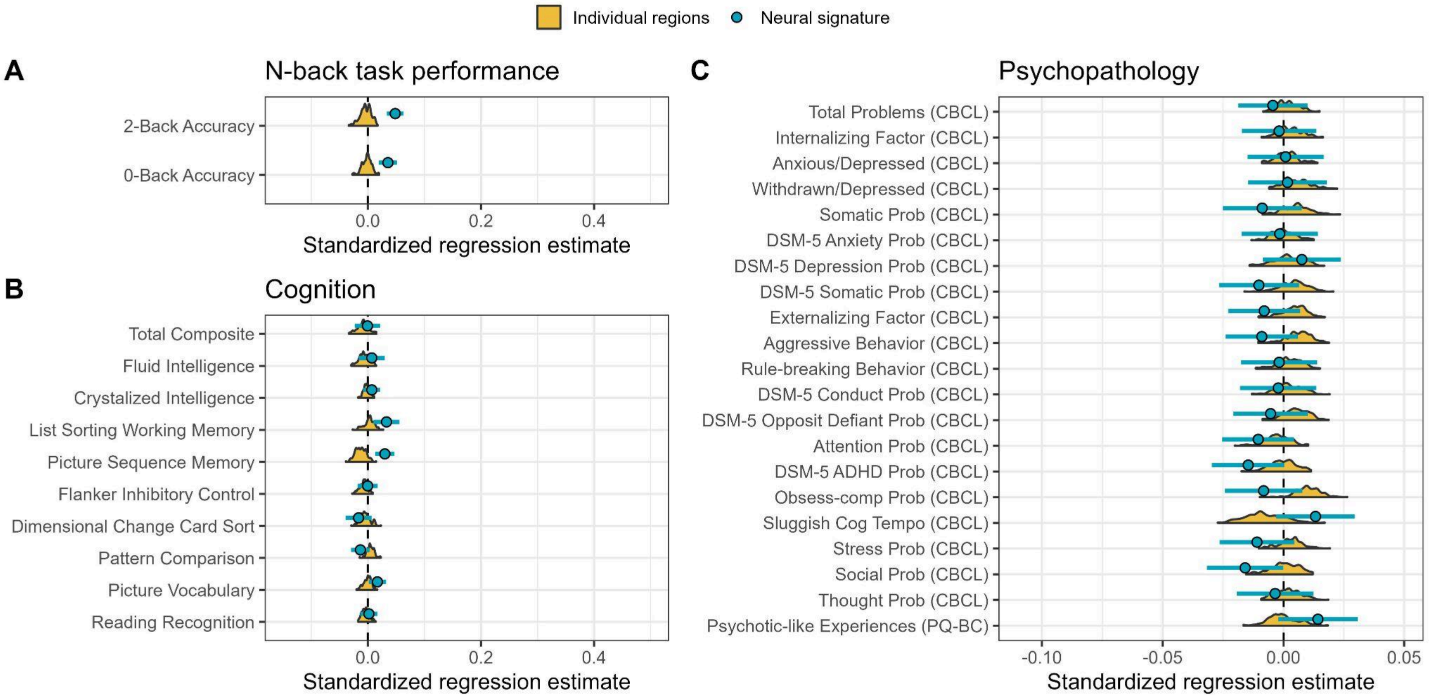


**Supplemental Figure 15**. **The Faces neural signature is not associated with individual differences in behavior, cognition, and psychopathology.** Associations of the faces neural signature with **A**) N-back task performance, **B**) NIH toolbox task performance and composite measures, and **C**) psychopathology. Shaded yellow represents the distribution of effects across all regions. Blue points reflect neural signature association estimates with 95% Confidence intervals (horizontal lines). Estimates reflect standardized regression coefficients.
